# A DDR2-targeted PET tracer images activated fibroblasts in early pulmonary fibrosis

**DOI:** 10.1101/2025.11.26.25341068

**Authors:** Xu Chen, Yunxin Lai, Penghui Yang, Guilin Li, Guodong Hu, Junhua Rao, Miao Shi, Qun Luo, Zhiying Chen, Jinquan Jiang, Lei Yang, Chuhui Gao, Ping Zhang, Haopeng Wang, Shizhen Qiu, Kun Wang, Xiaosong Ben, Jieqin Lv, Yuwang Cheng, Jiguo Liu, Jia Li, Pengcheng Ran, Jin Su

**Author notes:** Correspondence (Jin Su); (Jia Li); (Pengcheng Ran). These authors contributed equally (Xu Chen, Yunxin Lai, Penghui Yang, Guilin Li, Guodong Hu, Junhua Rao, Miao Shi).

## Abstract

Molecular imaging of activated fibroblasts holds promise for advancing the early detection and monitoring of idiopathic pulmonary fibrosis (IPF), a fatal lung disease with limited therapeutic options. Current imaging strategies primarily rely on fibroblast activation protein (FAP), but its functional ambiguity complicates the interpretation of signal intensity as a direct indicator of fibrotic activity. Here, we identify discoidin domain receptor 2 (DDR2), a collagen-binding receptor tyrosine kinase, as a superior and coherent target for imaging active fibroblasts in IPF. DDR2 was found to be significantly upregulated in IPF lung tissues, particularly in the early fibrotic phase, compared to FAP. Using a nanobody (1A12) with high affinity, specificity and stability, we developed a DDR2-targeted radiotracer, ^68^Ga-NOTA-1A12. In both mouse and non-human primate models of pulmonary fibrosis, we demonstrated that ^68^Ga-NOTA-1A12 PET/CT detected pathological changes earlier and with higher signal uptake than the established ^18^F-FAPI tracer, underscoring its potential for early diagnosis; moreover, total standardized uptake value of ^68^Ga-NOTA-1A12 correlated with fibrosis progression, providing a quantitative metric for disease evaluation. Notably, ^68^Ga-NOTA-1A12 PET/CT safely identified pulmonary fibrotic foci in a patient with interstitial lung disease. Our findings position DDR2-targeted PET/CT as a transformative tool for stratifying IPF patients, guiding anti-fibrotic therapies.

## Introduction

Idiopathic pulmonary fibrosis (IPF) is a lethal human lung disorder with limited treatment efficacy, partially due to a lack of reliable non-invasive techniques for early detection and patient stratification. Clinical trials remain dependent on functional declines, such as reductions in forced vital capacity (FVC), as enrollment criteria. This approach selects for patients who already have advanced, often irreversible fibrosis, rather than those in early pathogenic stage [1, 2]. This leads to substantial heterogeneity within trial cohorts, masking potential treatment effects in subgroups exhibiting early active fibrogenesis.

Molecular imaging with positron emission tomography (PET) represents a shift from visualizing structural remodeling with conventional CT to quantifying the dynamic disease biology in IPF, non-invasively assessing key processes such as activated fibroblast function and extracellular matrix (ECM) deposition [3–6]. Fibroblast activation protein (FAP) stands as the most clinically advanced target in this arena [7], with FAPI (fibroblast activation protein inhibitor) radiotracers providing robust tools for lesion visualization, burden mapping, and fibroblast quantitation in patients [8–10], and continued optimization of this technique is actively underway [11]. The field, however, is confronted by the paradoxical biology of FAP: while markedly upregulated in IPF [8, 12], its genetic ablation exacerbates experimental fibrosis, implicating it in ECM homeostasis. This functional duality thus complicates any straightforward interpretation of FAPI-PET signal intensity as a pure index of disease activity.

Discoidin domain receptor 2 (DDR2), a collagen-binding receptor tyrosine kinase, is emerging as a highly specific and mechanistically coherent target on activated fibroblasts in fibrosis. Given its dramatic upregulation in fibroblasts from IPF lungs [13], and the observation that its genetic ablation confers protection against fibrosis in murine models [14, 15], DDR2 is functionally implicated in disease progression. Mechanistically, it drives fibrogenesis in a biphasic manner: during early injury, it enhances TGF-β signaling in a ligand-independent manner, and upon collagen deposition, it transduces profibrotic signals through ERK and Akt pathways [14, 16]. This direct pro-fibrotic role, combined with its early involvement in disease pathogenesis, positions DDR2 as a compelling candidate for molecular imaging.

Here, we conduct a head-to-head comparative study of FAP and DDR2 as molecular targets for imaging IPF. Our strategy integrates single-cell RNA sequencing, flow cytometry, and histopathology to achieve a precise mapping of their expression across pulmonary cell types and niches. Central to this effort is the development of a novel DDR2-targeted nanobody PET/CT probe, ^68^Ga-NOTA-1A12, which we directly benchmark against the clinical-grade FAPI tracer ¹⁸F-FAPI-46 *in vivo*. This multi-modal framework enables a critical evaluation of both targets for visualizing active fibrogenesis, seeking to identify the superior candidate for early diagnosis and precision trial enrollment. Through both mouse and non-human primate models of pulmonary fibrosis, we demonstrate that ^68^Ga-NOTA-1A12 PET/CT is superior in early detection of pulmonary fibrosis. Importantly, ^68^Ga-NOTA-1A12 PET/CT safely traced the pulmonary pathological fibrotic sites in a human patient with interstitial lung disease. Our findings nominate DDR2 as a novel and promising PET target for diagnosing and monitoring disease progression in IPF, meriting subsequent clinical investigation.

## Results

### Expression Analysis of DDR2 and FAP in the Lung Tissue of Idiopathic Pulmonary Fibrosis (IPF) Patients

To identify the cell types expressing *DDR2* and *FAP* in fibrotic lung tissues, we first analyzed single-cell RNA sequencing (scRNA-seq) data from the public IPF dataset (GSE132771). The results showed that *DDR2* was primarily expressed in fibroblasts, myofibroblasts, PLIN2+ fibroblasts, and smooth muscle cells. In contrast, *FAP* expression was predominantly observed in fibroblasts and myofibroblasts, with overall expression levels lower than those of *DDR2* (Figure. 1a). To gain deeper insight, we performed re-clustering analysis of myofibroblast cell populations and found that *DDR2* was more highly enriched than *FAP* in IPF lung specimens (Figure. 1b). Analysis of another IPF dataset (GSE227136), stratified into control, less fibrotic and more fibrotic groups. Both *FAP* and *DDR2* were upregulated in IPF tissues compared to controls; interestingly, *DDR2* expression exceeded *FAP* in the less fibrotic group, whereas *FAP* was more enriched in the more fibrotic group (Figure. 1c). We next examined the expression of *TGFB1, DDR2, COL1A1,* and *COL1A2* across fibroblast, mesothelial, and myofibroblast cell clusters (GSE136831, Figure. 1d and Supplementary Figure. S1a). Correlation analysis revealed that *DDR2* expression was positively correlated with *TGFB1* (*R* = 0.33, *P* < 0.0001) but negatively correlated with *COL1A1* (*R* = -0.24, *P* < 0.0001) and *COL1A2* (*R* = - 0.24, *P* < 0.0001). Together, these findings demonstrate that both *DDR2* and *FAP* are significantly upregulated in IPF lung tissues. However, *DDR2* appears to be more prominent in the early fibrotic phase, suggesting its potential role as an early-stage molecular target in pulmonary fibrosis.

**Figure. 1.**
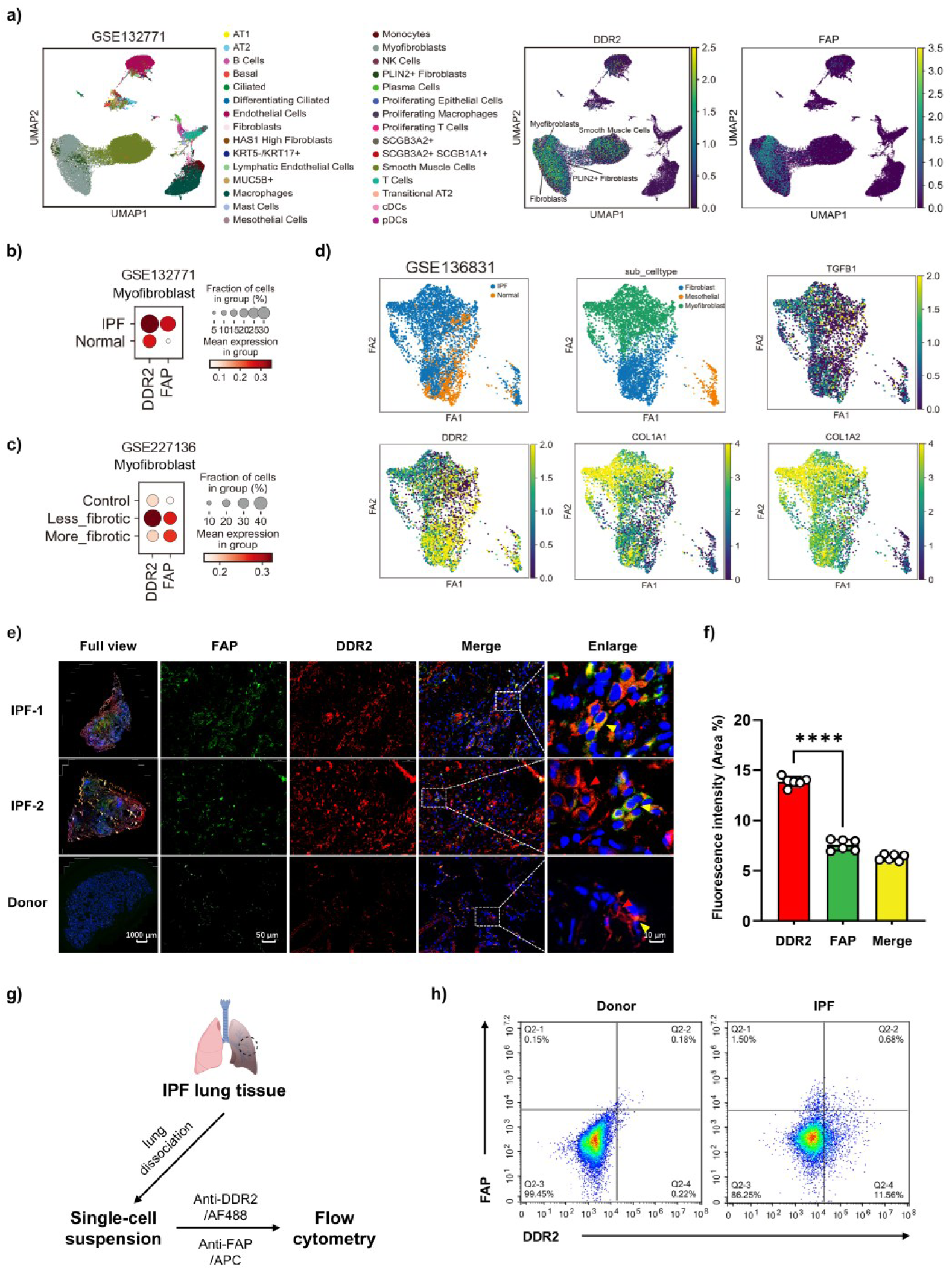
Expression profiles of *DDR2* and *FAP* in idiopathic pulmonary fibrosis (IPF) lung tissues. **a**, Single-cell RNA sequencing (scRNA-seq) analysis of the public IPF dataset (GSE132771) showing the distribution of cell populations (left) and UMAP plots indicating *DDR2* and *FAP* expression across different cell types (right). *DDR2* was primarily expressed in fibroblasts, myofibroblasts, PLIN2+ fibroblasts, and smooth muscle cells, while *FAP* was mainly restricted to fibroblasts and myofibroblasts with lower expression levels. **b**, Analysis of another IPF dataset (GSE136831) showing the expression of *TGFB1, DDR2, COL1A1,* and *COL1A2* in fibroblast and myofibroblast clusters. **c-d**, Quantitative comparison of *DDR2* and *FAP* expression in IPF and normal tissues (GSE132771) and in control, less fibrotic, and more fibrotic groups (GSE227136), revealing stage-dependent enrichment patterns. **e**, Representative immunofluorescence staining of DDR2 (red arrow) and FAP (green) in lung tissues from two IPF patients and one healthy donor. Nuclei were stained with DAPI (blue). DDR2 and FAP co-localization (yellow arrow) was observed mainly on the cell membrane. Scale bars: full view, 1000 μm; higher magnification, 50 μm; enlarged, 10 μm. **f**, Quantification of fluorescence-positive areas showing significantly higher DDR2 intensity compared to FAP (\*\*\*\**P* < 0.0001, *n* = 6, randomly selected fields from 2 independent IPF specimens). **g**, Schematic of the experimental workflow for single-cell suspension preparation and flow cytometry analysis using anti-DDR2/AF488 and anti-FAP/APC fluorescent antibodies. **h**, Flow cytometry analysis of donor and IPF lung tissues showing increased proportions of DDR2⁺ and FAP⁺ cells in IPF. All data are shown as mean ± SD; statistical significance was determined using one-way ANOVA. \*\*\*\**P* < 0.0001.

To further validate the single-cell sequencing results, we performed fluorescence staining and quantitative analysis using lung tissue samples from 2 IPF patients (Figure. 1e). Histological examination showed that alveolar structures were intact in the healthy donor group, while IPF patients exhibited significant fibrotic deposition with strong FAP and DDR2 fluorescence signals throughout the sections. Strong fluorescence signals of both FAP and DDR2 were observed throughout the fibrotic regions, with fluorescence areas significantly higher than those in donor control. Notably, both FAP and DDR2 were localized on the cell membrane (red arrows) and displayed clear co-localization (yellow arrows). Further analysis of co-localization revealed that FAP and DDR2 exhibited similar expression patterns in IPF lung tissues, with DDR2 showing stronger fluorescence intensity (Supplementary Figure. S1b). Quantitative analysis of fluorescence areas confirmed that the DDR2-positive area was significantly larger than that of FAP (DDR2, 13.86 ± 0.48% vs FAP, 7.50 ± 0.54%, *P* < 0.0001), and the co-localized area accounted for 6.37% of the total (Figure. 1f and Supplementary Figure. S1c). To further confirm the pathological findings, we prepared single-cell suspensions from IPF patient lung tissues and performed flow cytometry using fluorescence-conjugated antibodies anti-DDR2/AF488 and anti-FAP/APC (Figure. 1g). Compared with donor controls, the proportion of DDR2-positive cells was increased markedly from 0.22% to 11.56%, while FAP-positive cells increased from 0.15% to 1.50% in IPF tissues (Figure. 1h). Collectively, these results demonstrate that both DDR2 and FAP are significantly upregulated in the lung tissues of IPF patients, with DDR2 expression levels higher than FAP. These findings are consistent with the single-cell sequencing results and further confirm the distinct yet overlapping expression patterns of these two fibrosis-associated markers.

### Generation of DDR2-specific 1A12 Nanobody and ^68^Ga-NOTA-1A12 Probe

We developed DDR2-targeted nanobody 1A12, after conjugation with the bifunctional chelator *p*-SCN-Bn-NOTA, it was subsequently labeled with ^68^Ga to construct the ^68^Ga-1A12 radiotracer (details in Methods; Figure. 2a). Sodium dodecyl sulfate-polyacrylamide gel electrophoresis (SDS-PAGE) showed that both 1A12 and 1A12-NOTA had a molecular weight of approximately 15 kDa, with no extra bands (Figure. 2b). Further analysis by Ultra-performance liquid chromatography (UPLC) and time-of-flight mass spectrometry (TOF-MS) confirmed a purity of >95% for 1A12 and a molecular weight of 15,890 Da (Supplement Figure. 2a), consistent with typical nanobody characteristics. Biolayer Interferometry was performed to detect the affinity of 1A12 binding to soluble extracellular domain of DDR2 (Supplement Figure. 2b), and the calculated dissociation constant was 4.359e^-9^ M.

**Figure. 2.**
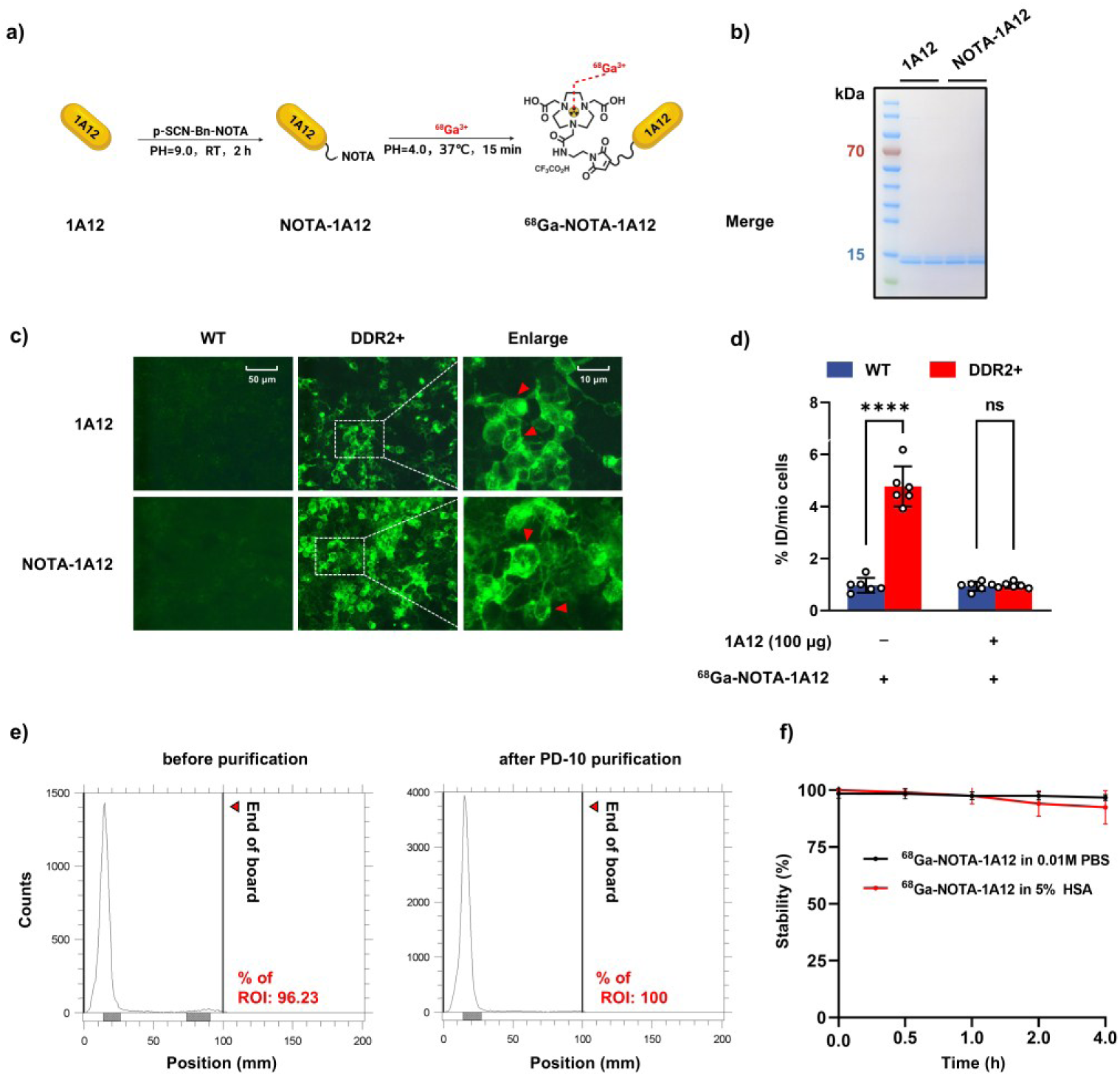
Characterization of the DDR2-targeted nanobody 1A12 and the ^68^Ga-NOTA-1A12 probe. **a**, Schematic illustration showing the conjugation of the DDR2-specific nanobody 1A12 with the bifunctional chelator *p*-SCN-Bn-NOTA, followed by radiolabeling with ^68^Ga to generate the ^68^Ga-NOTA-1A12 probe. **b**, SDS-PAGE analysis showing that both 1A12 and 1A12-NOTA exhibited a molecular weight of approximately 15 kDa with no additional nonspecific bands. **c**, Immunofluorescence staining of 293T and DDR2-overexpressing 293T (DDR2+) cells showing that both 1A12 and 1A12-NOTA localized mainly on the cell membrane (red arrows), whereas no fluorescence signal was detected in wild-type cells. **d**, *In vitro* binding and blocking assays demonstrating that ^68^Ga-1A12 exhibited high binding affinity to DDR2^+^ cells, which was effectively blocked by pre-incubation with unlabeled 1A12 (****P < 0.0001, ns = not significant; *n* = 6, one-way ANOVA). **e**, Radio–thin-layer chromatography (radio-TLC) showing radiochemical purities of 96.23% before and 100% after PD-10 purification. **f**, *In vitro* stability assay demonstrating that the radiochemical purity of ^68^Ga-1A12 remained above 90% after incubation in 0.01 M PBS or 5% human serum albumin (HSA) for up to 4 h, indicating good stability of the radiotracer *in vitro*. Scale bars: 50 μm; enlarged, 10 μm.

To evaluate the affinity and target specificity of 1A12 and 1A12-NOTA for DDR2, we used DDR2-overexpressing 293T cells. Flow cytometry revealed a distinct cell population in the DDR2+ group, with 82.08% being overexpression cells, and Western blot also confirmed high DDR2 expression in the DDR2+ group (Supplementary Figure. S2c-d). Since the 1A12 nanobody contains a His-tag at C terminus, we used a commercial anti-His-488 antibody as secondary labeling to label 1A12 and 1A121-NOTA. Fluorescence staining showed that both 1A12 and 1A12-NOTA localized mainly to the cell membrane surface (red arrows), while no fluorescence signal was detected in the wild-type (WT) group (Figure. 2c). Furthermore, to assess the probe’s specific binding to DDR2 *in vitro,* we performed a competitive blocking experiment. After constructing the ^68^Ga-1A12 radiotracer, we pre-incubate both WT and DDR2+ cells by 100 μg unlabeled 1A12 and find the binding of ^68^Ga-NOTA-1A12 to DDR2+ cells could be effectively blocked in the presence of 1A12 (Figure. 2d). These results indicate that both 1A12 and 1A12-NOTA have good affinity and specificity for DDR2, and that ^68^Ga-NOTA-1A12 retains high binding specificity to DDR2-overexpressing cells *in vitro*.

The ^68^Ga-NOTA-1A12 probe was manually radiolabeled (see Methods). Radio-TLC analysis showed that the radiochemical purity was 96.23% before and 100% after purification with a PD-10 desalting column (Figure. 2e). The approximate molar activity was 26.7∼48.3 GBq/μmol. The final injection solution was a clear, colorless liquid. After incubation in 0.01 M PBS or 5% human serum albumin, radio-TLC tests at 0.5, 1, 2, and 4 hours showed radiochemical purities above 90%, each presenting as a single peak, indicating that ^68^Ga-1A12 was stable *in vitro* within the time tested (Figure. 2f). In summary, we successfully constructed the ^68^Ga-NOTA-1A12 radiotracer and demonstrated its targeting specificity for DDR2 and its stability *in vitro*.

### Biodistribution, In Vivo Targeting, and Safety Assessment of the ^68^Ga-NOTA-1A12 Probe in Mice

To investigate the biodistribution of the ^68^Ga-NOTA-1A12 probe in mice, we conducted small-animal PET/CT imaging studies in C57BL/6 mice. At 40 min after injection, the ^68^Ga-NOTA-1A12 probe was mainly accumulated in the kidneys, with minor accumulation in the liver (Figure. 3a). To validate the PET/CT findings, biodistribution studies were conducted in C57BL/6 mice. The results showed that ^68^Ga-NOTA-1A12 uptake was predominantly in the kidneys, reaching a peak value of 26.86 ± 10.01 %ID/g at 1 h. Minor distribution was seen in the liver, kidney, bone, and lung tissues, which is consistent with the PET/CT imaging and aligns with the typical biodistribution pattern of nanobodies (Figure. 3b).

**Figure. 3.**
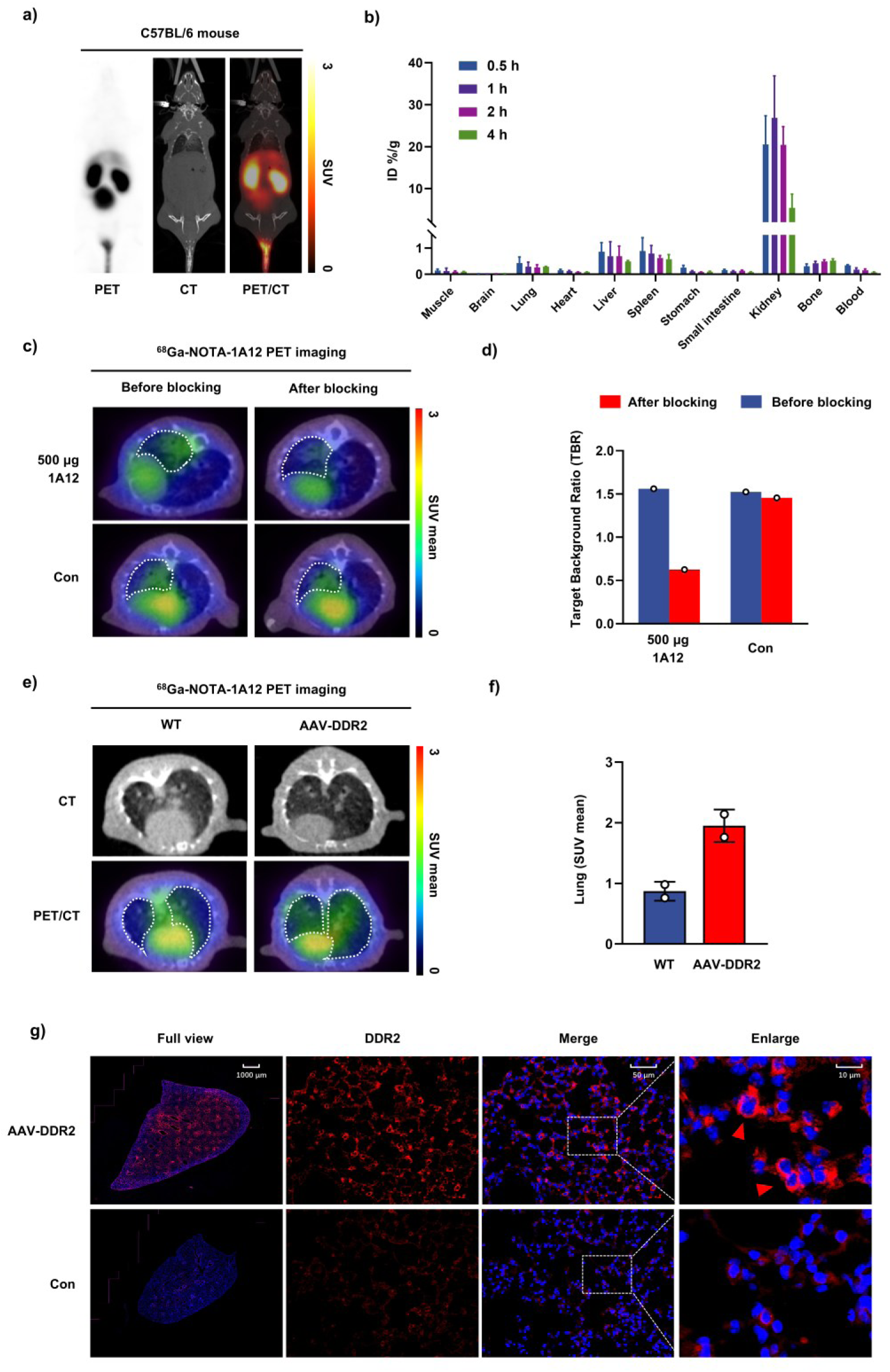
Biodistribution, *in vivo* targeting, and validation of the ^68^Ga-NOTA-1A12 probe in mice. **a**, Representative PET/CT images of C57BL/6 mice at 40 min after intravenous injection of ^68^Ga-NOTA-1A12 showing predominant accumulation in the kidneys and minor uptake in the liver, consistent with the renal clearance pattern typical of nanobodies. **b**, *Ex vivo* biodistribution analysis at 1 h post-injection showing the highest uptake of ^68^Ga-NOTA-1A12 in the kidneys (26.86 ± 10.01 %ID/g), with low levels in the liver, lung, bone, and other organs, consistent with the PET/CT results (*n* = 3). **c**, PET/CT images of unilateral bleomycin (BLM)-induced pulmonary fibrosis mice before and after blocking with unlabeled 1A12 (500 μg, 24 h interval). The second scan showed markedly reduced uptake in the fibrotic region after blocking. **d**, Quantitative analysis of the target-to-background ratio (TBR) showing a decrease from 1.56 to 0.625 in the blocking group, while remaining unchanged in the unblocked group. **e-f**, Schematic and PET/CT images of C57BL/6 mice after intratracheal administration of AAV-DDR2 demonstrating lung-specific DDR2 overexpression and significantly increased ^68^Ga-NOTA-1A12 uptake compared with wild-type (WT) mice, SUV mean = 1.95 ± 0.27 vs 0.87 ± 0.16). **g**, Representative immunofluorescence staining of lung sections from AAV-DDR2 and WT mice showing strong DDR2 expression the cell membrane (red, indicated by arrows) in the AAV-DDR2 group, confirming successful overexpression. Scale bars: full view, 1000 μm; higher magnification, 50 μm; enlarged, 10 μm.

To evaluate the *in vivo* specificity of ^68^Ga-NOTA-1A12 binding to DDR2, a competitive blocking experiment was performed. A unilateral intratracheal administration of bleomycin (BLM) induced pulmonary fibrosis mouse model was established as previously described (see details in Methods). On day 21, a distinct fibrotic region was observed in the left lung by micro-CT. The same BLM model mouse underwent two consecutive PET/CT scans (24 hours apart). For blocked group, 500 μg of unlabeled 1A12 was administered via tail vein injection after the first scan. The second PET/CT scan showed significantly reduced uptake in the fibrotic area of the lung. In contrast, unblocked group injected with PBS showed no obvious change in uptake within the fibrotic region. The target background ratio (TBR) decreased from 1.56 to 0.625 in the BLM-blocked group, while it remained unchanged in the BLM-unblocked group (Figure. 3c, d). Furthermore, to confirm specific targeting of DDR2 *in vivo*, we used adeno-associated virus (AAV)-DDR2 delivery system to achieve lung-specific DDR2 overexpression in C57BL/6 mice via intratracheal instillation, and PET/CT imaging and lung histopathological staining were used for mutual validation. PET/CT imaging with the ^68^Ga-NOTA-1A12 probe revealed a significantly higher uptake signal in AAV-DDR2 mice than in WT mice, with SUV mean values of 1.95 ± 0.27 vs 0.87 ± 0.16 (Figure. 3e, f). Immunofluorescence staining of lung tissues from the same mice showed strong DDR2 fluorescence signals in the AAV-DDR2 overexpression group, localized mainly to the cell membrane (red arrows), confirming successful DDR2 overexpression in the tissue (Figure. 3g). Combined imaging and pathological evaluation in the same DDR2-overexpressing mouse demonstrate that the ^68^Ga-NOTA-1A12 probe effectively recognizes DDR2 *in vivo*, indicating good targeting specificity.

We also assessed the safety profile of ^68^Ga-NOTA-1A12 probe. Mice were pre-injected with a dose exceeding 100 higher than the standard imaging dose. Body weight was monitored for 7 consecutive days, showing an increasing trend in both the ^68^Ga-NOTA-1A12 group and the control group, with no significant difference between them (Supplementary Figure. S3a). At the study endpoint, hematoxylin-eosin (H&E) staining of the heart, lungs, liver, kidneys, and spleen revealed normal tissue morphology in all examined organs (Supplementary Figure. S3b). Overall, we have characterized the biodistribution of the ^68^Ga-NOTA-1A12 probe and demonstrated its high *in vivo* targeting specificity for DDR2, and good biological safety, supporting its potential for further translational studies.

### PET/CT Imaging and Comparative Analysis of ^18^F-FAPI and ^68^Ga-1A12 Probes in Pulmonary Fibrosis Models

Since FAPI preferentially binds to human FAP, to better evaluate the imaging performance of the ^18^F-FAPI probe in murine pulmonary fibrosis, we generated a C57BL/6N-Fap^tm1(hFAP)/MCl^ mouse model with a humanized FAP sequence (construction details are provided in the Methods section). Subsequent experiments confirmed successful hFAP expression and its imaging applicability. First, Western blot confirmed successful hFAP expression in the lung tissues of hFAP knock-in (hFAP-KI) mice, with significantly elevated hFAP levels after BLM modeling (Supplementary Figure. S4a). Furthermore, we constructed an imaging probe hFAP-Cy7 by conjugating a Cy7 fluorescent dye to an anti-hFAP antibody. SDS-PAGE showed the probe’s molecular weight was approximately 30 kDa (Supplementary Figure. S4b-c). Using 293T cells overexpressing hFAP (hFAP+), both flow cytometry and *in vivo* imaging detected distinct cell population separation and fluorescent signal uptake in the hFAP+ group, indicating good *in vitro* targeting specificity of the probe for hFAP (Supplementary Figure. S4d-e). Finally, a unilateral pulmonary fibrosis model was established in hFAP-KI mice. Micro-CT scans and airway 3D modeling revealed significant fibrotic damage in the left lung, indicating successful modeling. *In vivo* fluorescence imaging showed strong fluorescent signals at the fibrotic site in hFAP-KI mice after BLM treatment, while no signal was detected in WT mice (Supplementary Figure. S4f). Correspondingly, PET/CT imaging of the same mice using the ^18^F-FAPI probe revealed high uptake in fibrotic regions of hFAP-KI mice but no significant uptake in WT BLM mice, which consistent with the fluorescence findings (Supplementary Figure. S4g). These results indicate that the ^18^F-FAPI probe exhibits superior uptake and good targeting capability in hFAP-KI pulmonary fibrosis mice, providing a valuable tool for subsequent PET/CT imaging studies of pulmonary fibrosis.

Subsequently, ^18^F-FAPI and ^68^Ga-NOTA-1A12 probes were used for sequential PET/CT imaging in the same batch of unilateral BLM-induced pulmonary fibrosis mice at days 3, 7, 14, 21, and 28. In the ^18^F-FAPI imaging group, the BLM-induced hFAP model mice showed no significant difference in signal uptake compared to the control group at day 3 (SUV mean, BLM 0.78 ± 0.31 vs Control 0.86 ± 0.25, *P* > 0.05). From day 7 to 28, the uptake gradually increased, with SUV mean values of 1.01 ± 0.29 at day 7, 1.09 ± 0.14 at day 14, 1.06 ± 0.34 at day 21, and peaking at 1.23 ± 0.26 at day 28, showing statistical significance compared to the control group. Interestingly, in the ^68^Ga-NOTA-1A12 imaging group, the lung fibrotic area in the BLM group already showed a statistically difference in uptake compared to the control group at day 3 (SUV mean, BLM 0.78 ± 0.31 vs Control 0.86 ± 0.25, *P* < 0.05). The uptake values gradually increased during the early stage of BLM-induced model (days 7 and 14), peaking at day 14 (SUV mean, 1.96 ± 0.39). However, during the later fibrotic stages (days 21 and 28), the uptake gradually decreased, resulting in a final SUV mean of 1.32 ± 0.30 at day 28 (Figure. 4a, b). Notably, the signal intensity of ^68^Ga-NOTA-1A12 at its peak was significantly higher than that of ^18^F-FAPI, although this observation requires more rigorous validation due to the different radionuclides used. To further compare the in vivo imaging performance of DDR2-targeted and FAP-targeted probes in the same animal, we designed a sequential dual-probe PET/CT imaging protocol in BLM-induced crab-eating macaque model of pulmonary fibrosis. The macaque was first imaged with ^18^F-FAPI on day 4 post-BLM instillation, followed by ^68^Ga-NOTA-1A12 PET/CT imaging on day 6 after allowing sufficient radioactive decay (Figure. 4c). Representative imaging data demonstrated that both probes visualized the fibrotic regions of the lung, whereas ^68^Ga-NOTA-1A12 displayed stronger and more localized uptake in fibrotic lesions. (Figure. 4d).

**Figure. 4.**
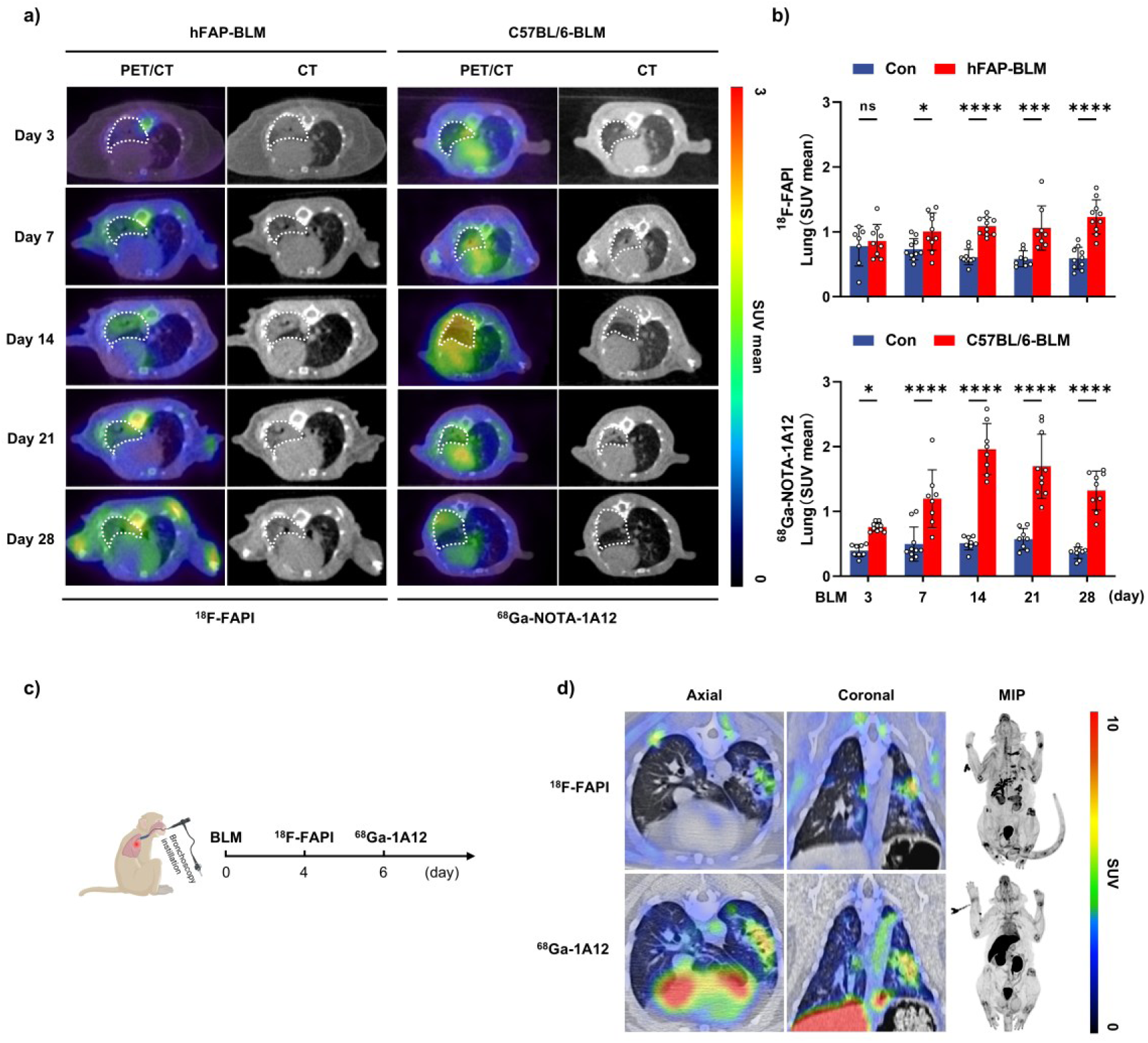
PET/CT imaging and comparative evaluation of ^68^Ga-NOTA-1A12 and ^18^F-FAPI probes in pulmonary fibrosis models. **a**, Representative longitudinal PET/CT and CT images of unilateral bleomycin (BLM)-induced pulmonary fibrosis in hFAP-knock-in (hFAP-BLM) mice imaged with ^18^F-FAPI and in wild-type C57BL/6-BLM mice imaged with ^68^Ga-NOTA-1A12 at days 3, 7, 14, 21, and 28 after modeling. White dashed lines delineate the fibrotic lung regions. **b**, Quantitative analysis of probe uptake in fibrotic lung regions expressed as SUV mean. Data are presented as mean ± SD (*n* = 8∼10 per group). Statistical significance was determined using one-way ANOVA. \**P* < 0.05, \*\**P* < 0.01, \*\*\**P* < 0.001, \*\*\*\**P* < 0.0001, ns: not significant. **c**, Schematic diagram of the sequential dual-probe PET/CT imaging protocol in the bleomycin-induced pulmonary fibrosis crab-eating macaque model. The macaque was first imaged with ^18^F-FAPI on day 4 post-BLM instillation, followed by ^68^Ga-NOTA-1A12 imaging on day 6 after sufficient radioactive decay. **d**, Representative axial, coronal, and maximum intensity projection (MIP) PET/CT images showing lung fibrotic regions with probe accumulation.

Based on the PET imaging data of the ^68^Ga-NOTA-1A12 probe in the unilateral pulmonary fibrosis model, we performed three-dimensional (3D) reconstruction of the lung regions and calculated the SUV total to provide a more comprehensive assessment of the probe’s overall uptake within the fibrotic lesions (Supplementary Figure. S5a). The PET 3D modeling results demonstrated that during the early stages of BLM-induced fibrosis (days 3, 7, and 14), the SUV total in the left lung fibrotic area showed a gradual increasing trend. The control group value was 0.26 ± 0.07, while the values at day 3 and day 7 were 0.61 ± 0.18 and 1.70 ± 0.56, respectively, peaking at 2.85 ± 0.48 on day 14. In the later fibrotic stages (days 21 and 28), the SUV total gradually decreased to 1.83 ± 0.56 and 1.42 ± 0.23, respectively (Supplementary Figure. S5b). This trend was consistent with the pattern observed in the previous SUV mean analysis. In summary, compared to the conventional SUV mean and SUV max calculations, the SUV total value may offer a more comprehensive metric for evaluating the total radiotracer uptake throughout the fibrotic lung lesions.

Together, these results demonstrate that the DDR2-targeted ^68^Ga-NOTA-1A12 probe provides earlier and stronger uptake in fibrotic lung lesions than the FAP-targeted ^18^F-FAPI probe across both mouse and non-human primate models. The hFAP-KI system confirmed the specificity of FAPI imaging, while longitudinal and 3D quantitative analyses showed that ^68^Ga-NOTA-1A12 is more sensitive to early fibrotic changes and offers a more comprehensive assessment of lesion burden. Overall, ^68^Ga-NOTA-1A12 shows clear advantages for early detection and monitoring of pulmonary fibrosis.

### Expression Analysis of DDR2 and FAP in the Lung Tissue of Unilateral Pulmonary Fibrosis Mouse Model

To verify the specificity and applicability of the anti-mouse FAP antibody prior to lung tissue staining, we first validated its performance in an *in vitro* overexpression system. MC38 cells stably overexpressing murine FAP (MC38-mFAP) and wild-type MC38 cells were subjected to immunofluorescence staining (Supplementary Figure. S6). MC38-mFAP cells exhibited strong and membrane-localized green fluorescence signals corresponding to FAP, whereas MC38 wild-type cells showed only minimal background staining. These results confirm that the anti-mouse FAP antibody specifically recognizes FAP and is suitable for subsequent fluorescence staining experiments in mouse lung tissues. To further validate the PET/CT imaging findings, we performed fluorescence staining and quantitative pathological analysis of lung tissues from the unilateral pulmonary fibrosis mouse model at days 3, 7, 14, 21, and 28 after BLM treatment (Figure. 5a). Strong fluorescence signals of both FAP and DDR2 were observed in the fibrotic regions (Figure. 5b). The FAP (green) fluorescence area showed a gradual increase over time, from 3.68 ± 0.70% (Day 3) to a peak of 24.16 ± 3.48% (Day 28). In contrast, the DDR2 (red) fluorescence area increased during the early stages of fibrosis, from 9.92 ± 2.87% (Day 3) to a peak of 30.68 ± 4.75% (Day 14). However, during the later stages (Day 14 to Day 28), the DDR2 fluorescence area gradually decreased to 14.74 ± 3.68% (Day 28). Notably, from BLM-days 3 to 14, the DDR2-positive area was significantly larger than that of FAP (*P* < 0.0001). As FAP expression continued to rise, the difference between the two became insignificant at day 21 (FAP: 20.40 ± 3.56% vs DDR2: 21.07 ± 4.02%, *P* > 0.05). By day 28, the FAP-positive area became significantly larger than the DDR2-positive area (FAP, 24.16 ± 3.48% vs DDR2, 14.74 ± 3.68%, *P* < 0.0001). We also analyzed the co-localization of FAP and DDR2 signals (yellow) (Figure. 5c). The co-localized area peaked at 16.30 ± 3.66% on day 21, which interestingly corresponds to the typical endpoint of the BLM-induced fibrosis model. These findings suggest that although both FAP and DDR2 serve as specific markers of fibroblast activation, they may reflect different underlying mechanisms and fibrotic states. In summary, the expression patterns of FAP and DDR2 in the fibrotic lung regions align with the PET/CT imaging results obtained using the ^18^F-FAPI and ^68^Ga-1A12 probes.

**Figure. 5.**
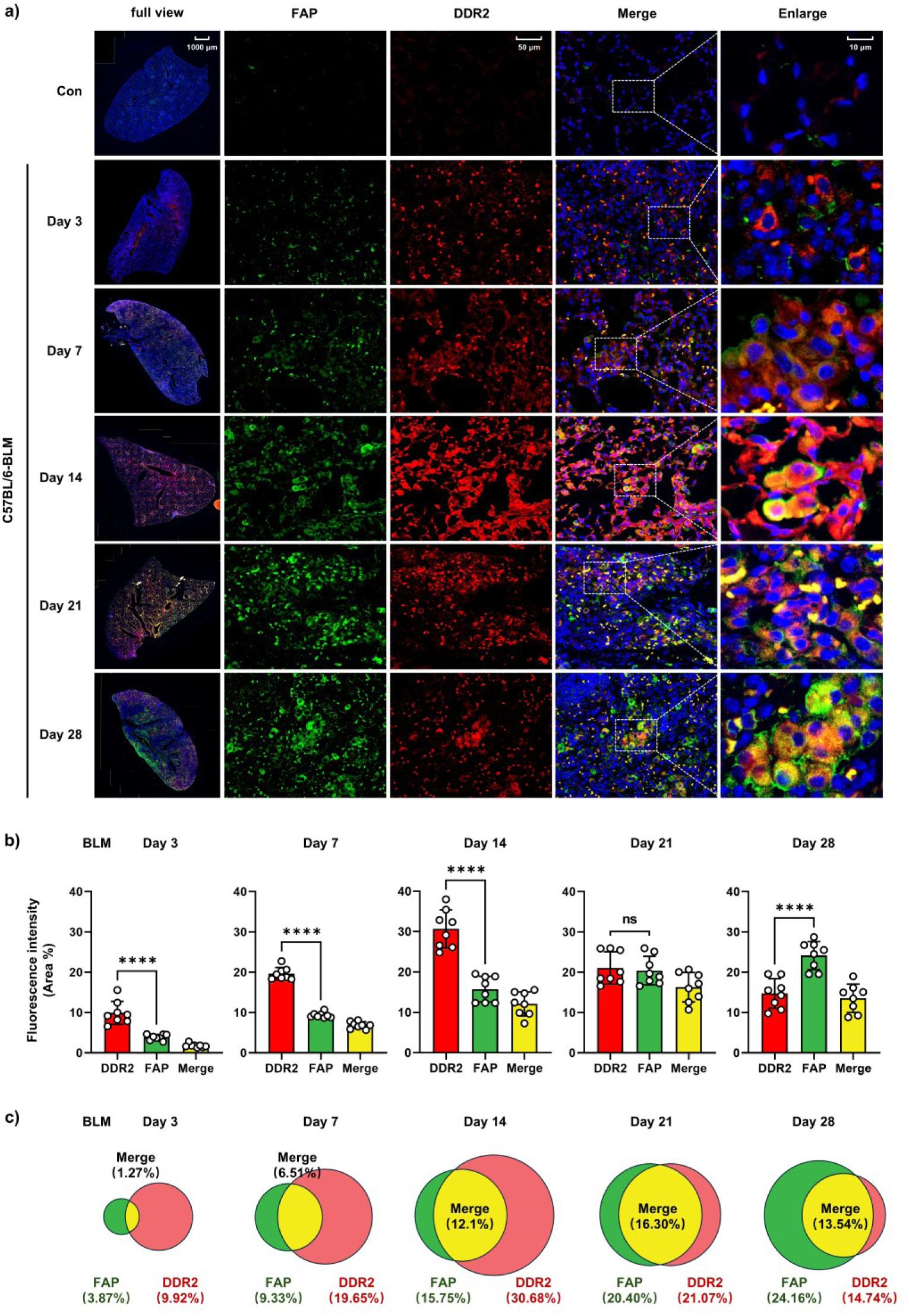
Temporal expression patterns of DDR2 and FAP in lung tissues of unilateral BLM-induced pulmonary fibrosis mice. **a,** Representative immunofluorescence staining of lung sections from control and BLM-treated C57BL/6 mice at days 3, 7, 14, 21, and 28. Full-view images show whole-lung distribution of fluorescence signals, while higher magnification fields reveal localized expression of FAP (green) and DDR2 (red) within fibrotic regions. Nuclei were counterstained with DAPI (blue). Progressive increases in FAP and DDR2 signals were observed following BLM instillation, with DDR2 showing strong early-stage expression (Days 3∼14) and FAP exhibiting a sustained rise that peaked at day 28. Enlarged panels highlight membrane-associated signals and areas of co-localization (yellow). Scale bars: full view, 1000 μm; higher magnification, 50 μm; enlarged, 10 μm. **b,** Quantification of fluorescence-positive areas for DDR2, FAP, and their merged regions across time points (*n* = 6 mice per group, biological replicates; values represent the mean fluorescence area per mouse averaged from multiple randomly selected fields). DDR2 expression peaked at day 14 and then declined, whereas FAP expression increased continuously through day 28. Statistical analysis: one-way ANOVA, \*\*\*\**P* < 0.0001; ns, not significant. **c,** Venn-diagram style visualization of DDR2-positive, FAP-positive, and co-localized (Merge) fluorescence areas at each time point. Co-localized areas progressively increased and reached their maximum at day 21.

### PET/CT Imaging in a Patient with Interstitial Lung Disease

To further explore the translational potential of the DDR2-targeted tracer, we conducted a preliminary clinical PET/CT evaluation in a patient with clinically diagnosed interstitial lung disease (ILD). whole-body maximum intensity projection (MIP) PET images after ^68^Ga-1A12 (3.0MBq/kg) injection showed prominent tracer accumulation in the kidneys and urinary bladder, with only mild physiological uptake in the liver and heart (Figure. 6a). Subsequent axial, coronal, and sagittal fused PET/CT images demonstrated that ^68^Ga-1A12 accumulation was predominantly localized within radiographically abnormal pulmonary areas, particularly those exhibiting reticulation, interlobular septal thickening, and subpleural fibrotic changes on CT (Figure. 6b). Regions with relatively preserved lung parenchyma displayed minimal tracer uptake, suggesting a strong association between ^68^Ga-1A12 signal intensity and fibrotic remodeling.

**Figure. 6.**
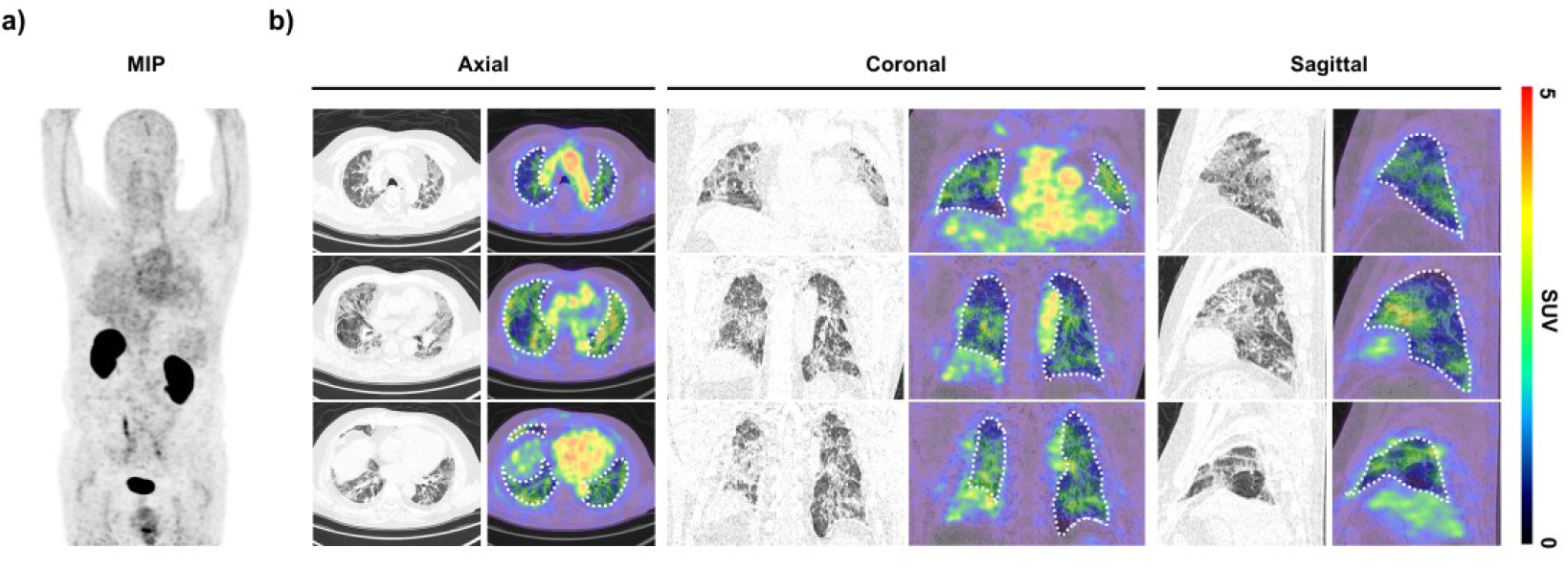
Whole-body PET/CT imaging of a patient with interstitial lung disease using ^68^Ga-NOTA-1A12. **a,** Maximum intensity projection (MIP) PET image acquired 1 h after intravenous administration of ^68^Ga-NOTA-1A12, showing prominent physiological uptake in the kidneys and urinary bladder, with mild uptake observed in the liver and heart. **b,** Representative axial, coronal and sagittal PET/CT fusion images. Dotted lines outline the lung fields. A distinct area of increased tracer uptake is observed in the fibrotic region of the lung, indicating enhanced DDR2 expression in diseased tissue. This focal uptake markedly exceeds the background activity of normal lung parenchyma.

Importantly, the lesion-to-background contrast was high across all imaging planes, allowing clear delineation of abnormal fibrotic regions. The spatial distribution of tracer uptake showed a pattern consistent with the known heterogeneity of ILD, including patchy subpleural involvement and basilar predominance. These findings are aligned with the preclinical animal imaging results, in which DDR2-targeted radiotracers selectively accumulated in fibrotic regions and reflected dynamic fibroblast activation. Notably, no adverse events were observed within 24 h of ⁶⁸Ga-1A12 injection, indicating a favorable initial safety profile. Collectively, this first-in-human observation supports the feasibility of DDR2-targeted PET/CT imaging in lung fibrosis and highlights the potential clinical utility of ⁶⁸Ga-1A12 for noninvasive assessment of fibroblast activation in ILD.

## Discussion

This study provides a definitive comparative evaluation of fibroblast-associated molecular imaging targets in IPF, establishing DDR2 as a superior biomarker to FAP. Through integrated multimodal data, we demonstrate that a novel DDR2-PET probe outperforms the clinical-stage FAPI tracer in both sensitivity to active fibrogenesis and signal robustness in early disease. This divergence stems from fundamental biological differences. DDR2 acts as an early-response sentinel, generating a PET signal characterized by early emergence, a rapid peak, and subsequent attenuation with collagen maturation, thereby confirming its specificity for active disease phases. In contrast, FAP participates in late-stage remodeling, exhibiting only a gradual signal increase. The reduction of DDR2-PET signal in collagen-dense lesions thus reflects not a technical limitation but a biologically meaningful delineation of a treatable window.

Clinically, a positive DDR2-PET scan selects ideal candidates for anti-fibrotics targeting initiation pathways, while signal absence in structurally abnormal areas suggests inactive scar, potentially sparing patients from futile interventions. FAPI-PET, by comparison, visualizes advanced fibrotic niches likely requiring ECM-modifying strategies. Longitudinal DDR2-PET imaging further offers unique pharmacodynamic insight, enabling direct assessment of treatment efficacy for early-pathway inhibitors, whereas FAP signal dynamics necessitate cautious interpretation. Together, this noninvasive functional mapping supports real-time, mechanism-guided therapeutic sequencing in IPF.

Looking beyond IPF, the implications of our DDR2-PET probe extend strategically into the burgeoning field of DDR2-targeted therapies across a spectrum of diseases. DDR2 overexpression or pathogenic activation has been implicated in several malignancies, where it drives tumor progression, invasion, and stromal fibrosis [17–20]. As DDR2-specific inhibitors advance in preclinical and clinical development [21–23], a central challenge will be the precise identification of patients with DDR2-dependent pathologies. Our DDR2-PET platform directly addresses this need, positioning it as an ideal companion diagnostic to non-invasively quantify in vivo DDR2 expression before treatment initiation.

In summary, this work moves beyond the mere description of a new imaging target to deliver a rigorous, head-to-head comparison that resolves a central question in IPF and fibrosis research. We present compelling evidence that DDR2, by virtue of its specific and integral role in the fibrotic cascade, offsets FAP for molecular imaging applications. The DDR2-PET probe developed herein is not merely a new diagnostic tool but a potential catalyst for a new era of preemptive, biomarker-driven clinical trials in IPF, ultimately aiming to intercept fibrosis at its earliest.

## Data Availability

All data produced in the present study are available upon reasonable request to the authors

## Acknowledgements

We would like to thank Dr. Junhui Yang (Guangzhou National Laboratory) for her integrative scRNA-seq data analysis to identify biomarkers for pulmonary fibrosis. The graphical abstract was generated with BioRender software (www.biorender.com/).

## Method

### Human Lung Tissue Samples

Human lung tissue samples were obtained with approval from the Ethics Committee of the First Affiliated Hospital of Guangzhou Medical University. Written informed consent for tissue collection was obtained from all participants or their legal guardians. Lung specimens from patients with idiopathic pulmonary fibrosis (IPF) were collected during lung transplantation or diagnostic lung biopsy procedures. The diagnosis of IPF was established by the patients’ attending pulmonologists in accordance with the guidelines of the American Thoracic Society and the European Respiratory Society. Healthy lung tissues used as controls were collected from brain-dead organ donors without evidence of pulmonary disease.

### Single-cell Analysis

Publicly available scRNA-seq datasets of idiopathic pulmonary fibrosis (IPF) were obtained from the Gene Expression Omnibus (GEO) database, including GSE132771, GSE227136, and GSE136831 to characterize DDR2 and FAP expression patterns. Raw sequencing data were processed using Cellranger (v6.0) with GRCh38 as the reference genome to generate gene-cell count matrices. Data preprocessing, quality control, normalization, dimensionality reduction, clustering, and visualization were performed using Seurat (v4.0) in R (v4.1.2). Cells with fewer than 200 detected genes, more than 10% mitochondrial gene content, or identified as doublets by DoubletFinder were excluded. Genes detected in fewer than 50 cells were removed. Cell-type annotation was conducted based on canonical marker genes reported in previous studies. For comparative analysis, cell populations of interest were re-clustered, and differential expression analysis was performed using the Wilcoxon rank-sum test. Correlation analyses between DDR2 and fibrosis-related genes were conducted using Pearson’s correlation coefficient. Data visualization, including UMAP plots, box plots, and scatter plots, was generated using Seurat and ggplot2 packages.

### Production and Purification of DDR2-Targeting Nanobody 1A12

The DDR2-targeting nanobody named 1A12 containing a C-terminal His tag was produced in *Escherichia coli (E. coli)* as previously described (PMID: 33563286, 34552091). Briefly, the DNA sequence encoding the anti-DDR2 single-domain nanobody was screened using phage display technology and subcloned into the pET-28a(+) expression vector fused with a His tag for bacterial expression. The recombinant plasmid was transformed into *E. coli* BL21 (DE3) competent cells, and protein expression was induced with isopropyl β-D-1-thiogalactopyranoside (IPTG; MedChemExpress) at 20°C overnight in a shaking culture. Bacterial cells were harvested and lysed by ultrasonic disruption to obtain the cytoplasmic extract. The nanobody was purified from the supernatant using Nickel-Nitrilotriacetic Acid (Ni-NTA; GenScript) affinity chromatography and then buffer-exchanged into phosphate-buffered saline (PBS; Gibco) for subsequent use. The purity of the nanobody was assessed by sodium dodecyl sulfate-polyacrylamide gel electrophoresis (SDS-PAGE; ZHHC Biotechnology Co., Ltd, Shaanxi, China); under optimal conditions, a single distinct band of approximately 15 kDa was observed after staining with Coomassie Brilliant Blue. The purified product was stored at -20°C until further use.

#### UPLC–QTof Analysis of Nanobody Molecular Weight

The molecular weight and purity of the 1A12 nanobody were analyzed using an ultra-performance liquid chromatography coupled with high-resolution quadrupole time-of-flight mass spectrometer (UPLC-QTof) system (ACQUITY UPLC I-Class PLUS/Xevo G2-XS QTof; Waters, USA). Chromatographic separation was performed on an ACQUITY UPLC Protein BEH C4 column (2.1 ×50 mm, 1.7 μm; Waters), with the column temperature set at 80°C and autosampler at 10°C. A 10 μL aliquot of sample (500 ng) was injected. The mobile phases consisted of solvent A (0.1% formic acid in water) and solvent B (acetonitrile), using the following gradient: 10-20% B from 1-4 min, 20-30% B from 4-13 min, and increased to 90% B at 15 min, with a flow rate of 0.3 mL/min. UV absorbance was monitored at 280 nm. Mass spectrometry was performed in positive ion mode with a resolving power of 30,000, micro-scan of 10, and an m/z acquisition range of 400–5000. Raw spectra were processed using MaxEnt1-based deconvolution algorithms (Waters MassLynx software, Waters, USA) to obtain the zero-charge molecular mass profile of the nanobody for subsequent purity and identity confirmation.

#### DDR2-Targeting Nanobody 1A12 Affinity Assay

A549 cells were cultured in Dulbecco’s modified Eagle’s medium (DMEM; Gibco) supplemented with 10% fetal bovine serum (FBS; Gibco) and 1% penicillin-streptomycin (P/S; Gibco). To generate DDR2-overexpressing cells, hDDR2-expressing adenovirus (OBiO Technology, Shanghai, China) was added to A549 cells when they reached approximately 90% confluence and incubated for 24 h. The binding affinity of the DDR2-targeting nanobody (sdNb) 1A12 was evaluated by immunocytochemical staining. A549 and A549-DDR2 cells were seeded in 24-well plates at a density of 1×10⁵ cells per well and cultured for 24 h. Cells were then fixed with 4% paraformaldehyde for 30 min, permeabilized with 0.5% Triton X-100 for 10 min, and blocked with 5% bovine serum albumin (BSA; Aladdin) for 1 h. Subsequently, the cells were incubated with 1 μg of sdNb, followed by incubation with Alexa Fluor 488-conjugated anti-His tag IgG (MBL, D291-A48; 1:400 dilution in 1% BSA) for 1 h at room temperature. Cell nuclei were counterstained with 4’,6-diamidino-2-phenylindole (DAPI; Beyotime). Finally, fluorescence images were acquired using an inverted fluorescence microscope (ZEISS, Germany).

#### Fluorescent Labeling of hFAP Antibody

To generate fluorescently labeled hFAP antibody, the antibody was conjugated with an N-hydroxysuccinimide (NHS) ester-activated fluorophore, Cyanine 7 (Cy7; MCE), via reaction with lysine amino groups. Briefly, the hFAP antibody was dialyzed in PBS to final concentration of 2 mg/mL and then incubated with the Cy7 fluorophore dissolved in DMSO at a molar ratio of 1:5 for 2 h at room temperature. After the reaction, unbound fluorophore molecules were removed using a 3 kDa ultrafiltration tube (Millipore, USA).

#### Radiolabeling of 1A12 Nanobody with ^68^Ga

The DDR2-targeting nanobody 1A12 was conjugated with the bifunctional chelator 2-S-(4-isothiocyanatobenzyl)-1,4,7-triazacyclononane-1,4,7-triacetic acid (*p*-SCN-Bn-NOTA; Macrocyclics, USA) for subsequent ⁶⁸Ga labeling. Briefly, 2 mg of 1A12 dissolved in PBS were adjusted to pH 9.0 with 0.1 M Na₂CO₃ buffer. *p*-SCN-Bn-NOTA dissolved in DMSO was then added at a molar ratio of 10:1 (chelator: nanobody) under gentle vortexing. The reaction mixture was incubated at room temperature for 2 h, after which the NOTA-conjugated nanobody was purified using a PD-10 desalting column (Cytiva, USA) and concentrated with a 3 kDa ultrafiltration tube (Millipore, USA). The purified conjugate was stored at 4 °C for subsequent radiolabeling. For radiolabeling, a ^68^Ge/^68^Ga generator (New Radiomedicine Technology, China) was eluted with 2 mL of 0.1 M HCl to obtain freshly eluted ^68^Ga. The eluate was then adjusted to pH 4.0 with 3 M sodium acetate buffer (Macklin, China). The NOTA-conjugated nanobody was added to 2 mL of the ^68^Ga solution (pH 4.0), and the mixture was incubated at 37°C with gentle shaking (600 rpm) for 15 min. The resulting radiolabeled product, ^68^Ga-1A12, was purified using a pre-equilibrated PD-10 column with sterile PBS as the mobile phase. Radiochemical purity and labeling efficiency were analyzed using instant thin-layer chromatography (ITLC; Eckert & Ziegler, Germany).

#### Preparation of ^18^F-FAPI-46

The ^18^F-FAPI-46 tracer was obtained from Guangdong Gosun Cyclotron Medicine Co., Ltd. (Guangzhou, China). Radiolabeling was performed on-site using a medical cyclotron to produce ^18^F-fluoride, followed by automated synthesis of ^18^F-FAPI-46 according to the manufacturer’s standardized protocol. After radiochemical processing and purification, the final product was formulated and passed through a sterile filter, and the radioactive solvent was replaced with sterile phosphate-buffered saline (PBS) to obtain the injectable ^18^F-FAPI-46 preparation suitable for *in vivo* imaging. Radiochemical purity, pH, sterility, and endotoxin levels all met the quality control requirements for clinical PET imaging.

#### in vitro Stability Assay

*In vitro* stability of ^68^Ga-1A12 was evaluated by incubating the radiolabeled nanobody in either saline or 5% human serum albumin (HAS, ab205808, abcam) at 37°C for up to 4 h. At predetermined time points, samples were collected and analyzed for radiochemical purity using radio-instant thin layer chromatography (radio-ITLC).

#### Tissue Biodistribution Study

C57BL/6 mice were euthanized 1 h after intravenous injection of approximately 0.74 MBq of the ^68^Ga-1A12 probe. Major organs, including the heart, liver, lungs, kidneys, spleen, stomach, bone, small intestine, muscle, blood, and brain, were rapidly harvested, rinsed, and weighed. The radioactivity in each tissue was measured using a γ-counter (PerkinElmer, USA). The results were expressed as the percentage of injected dose per gram of tissue (%ID/g) for each organ. For the lungs, radioactivity was additionally calculated and presented as the percentage of injected dose per lung (%ID/lung).

### Cell Culture and Construction of Stable Overexpression Cell Lines

HEK293T cells were cultured in Dulbecco’s modified Eagle’s medium (DMEM; Gibco) supplemented with 10% fetal bovine serum (FBS; Gibco) and 1% penicillin-streptomycin (Gibco) at 37°C in a humidified atmosphere containing 5% CO₂. To generate stable overexpression models, lentiviral vectors carrying human DDR2 or hFAP complementary DNA (IGE Biotechnology, Guangzhou, China) were constructed and co-transfected with packaging plasmids psPAX2 and pMD2.G into 293T cells using Lipofectamine 3000 (Invitrogen). The culture supernatants were collected at multiple time points after transfection and concentrated by ultrafiltration (Millipore) to obtain lentiviral particles. When target 293T cells reached approximately 50% confluence, they were infected with the virus-containing medium for 48 h and subsequently cultured in complete medium. Stable clones were selected with puromycin (2 µg/mL, MedChemExpress, USA) for about two weeks to establish the 293T-DDR2+ and 293T-hFAP+ cell lines. Wild-type 293T cells served as controls. The expression of DDR2 and hFAP in the stable lines was verified by Western blotting, immunofluorescence staining, and flow cytometry before subsequent *in vitro* and imaging experiments. Similarly, MC38-mFAP cells were generated using the same lentiviral transduction procedure. Briefly, MC38 murine colon carcinoma cells were cultured under identical conditions and infected with lentivirus encoding mouse FAP, followed by puromycin selection to obtain a stable MC38-mFAP cell line.

### 68Ga-NOTA-1A12 Cell Binding Assay

For the cell binding assay, 293T-DDR2+ overexpression cells and wild-type 293T cells were seeded in 24-well plates at a density of 2×10⁵ cells per well and cultured for 24 h. At the start of the experiment, the culture medium was removed and replaced with 500 μL of DMEM (Gibco, USA) containing approximately 74 kBq of ^68^Ga-1A12. Cells were incubated at 37°C in 5% CO₂ for 60 min. For the blocking control, 100 μg of unlabeled 1A12 nanobody was added simultaneously to the wells. After incubation, the medium was discarded, and cells were washed twice with 500 μL of cold PBS. Subsequently, 300 μL of 1 M NaOH (Macklin, China) was added to lyse the cells for 5 min, after which the lysates were collected and measured using a γ-counter (PerkinElmer, USA). Data were analyzed using Prism 8.0 software (GraphPad Software, USA) and expressed as the percentage of injected dose per million cells (%ID/10⁶ cells).

#### Unilateral Pulmonary Fibrosis Animal Model

All animal protocols used in this study conformed to the requirements of the Animal Welfare Act and were approved by the Animal Care and Use Committee of Guangzhou Medical University. BLM-induced mice model of pulmonary fibrosis: Male C57BL/6J mice (6-8 weeks) were purchased from Hunan SJA Laboratory Animal Co., Ltd (Changsha, Hunan, China) and housed in the specific pathogen-free (SPF) barrier area. To induce unilateral pulmonary damage, 45 μL bleomycin sulfate (BLM, MedChemExpress, HY-17656A) dissolved in saline at the concentration of 0.75 mg/mL was injected into the left lung lobes via intratracheal instillation under anesthesia by pentobarbital sodium.

BLM-induced crab-eating macaque model of pulmonary fibrosis: Adult male crab-eating macaque weighing between 9-10 kg were provided by Institute of Zoology, Guangdong Academy of Sciences and housed individually with a 12 h light/dark cycle. The anesthetized crab-eating macaque were administered approximately 10 mL bleomycin at the concentration of 0.75 mg/mL into the left lung lobes through endotracheal intubation with the assistance of fiberoptic bronchoscopy.

#### Generation and Housing of Humanized FAP Knock-In Mice

C57BL/6N-Fap^tm1(hFAP)/MCl^ mice were generated by Guangzhou Mingceler Biotech Co., Ltd. (Guangzhou, China), and the construction strategy is illustrated below. A FAP-targeting plasmid containing the human FAP coding sequence (CDS) was constructed and introduced into embryonic stem (ES) cells by electroporation. Homozygous mutant ES cell lines were obtained through homologous recombination, and homozygous mice were subsequently generated using the tetraploid embryo complementation method. The inserted hFAP CDS was verified by PCR amplification using the following primers: forward 5’-GTAAAAATCGTATTTGGAGTTGCCACC-3’ and reverse 5’-CTGCAAACATACTCGTTCATCAGTAACC-3’. A 909 bp PCR product was obtained and confirmed to contain the correct human FAP sequence by Sanger sequencing. All mice were maintained under specific pathogen-free (SPF) conditions in individually ventilated plastic cages (2-3 mice per cage), with sentinel mice periodically monitored for pathogens throughout the study. The housing environment was maintained at 23.5 ± 2.5 °C and 52.5 ± 12.5% relative humidity under a 14-h light/10-h dark cycle. Mice had ad libitum access to standard laboratory chow and autoclaved water. All animal experiments were conducted in accordance with institutional and national guidelines and were approved by the Institutional Animal Care and Use Committee (IACUC) of Guangzhou Mingceler Biotech Co., Ltd.

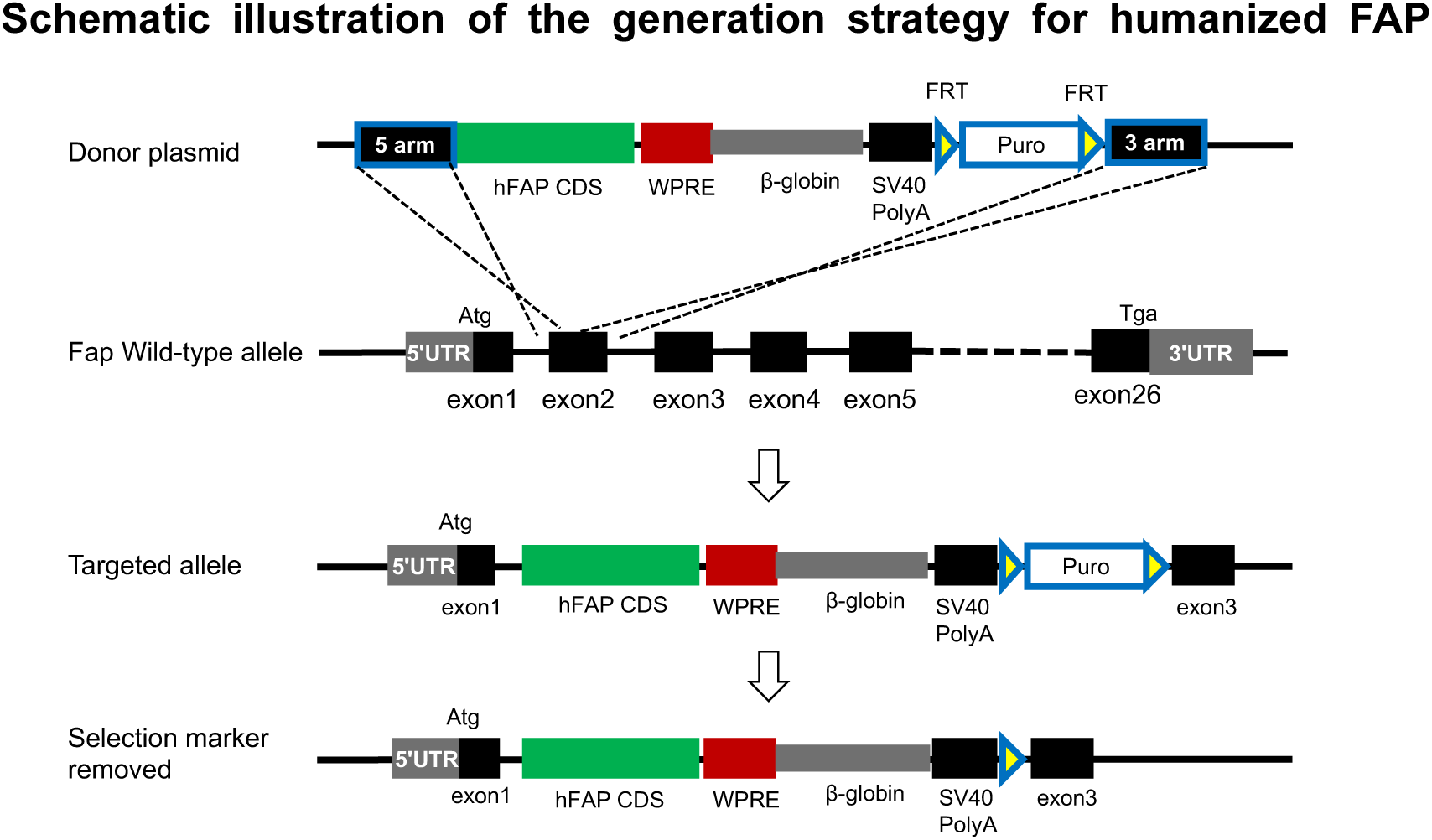
knock-in (hFAP-KI) mice. A targeting construct containing the human FAP coding sequence (CDS) flanked by 5′ and 3′ homology arms and a puromycin selection cassette was introduced into mouse ES cells by electroporation. Homologous recombination replaced the endogenous mouse Fap CDS with the human FAP CDS, and correctly targeted ES clones were identified through antibiotic selection and PCR screening. Homozygous knock-in mice were subsequently generated via tetraploid embryo complementation. Successful insertion of the hFAP CDS was verified by PCR amplification of a 909-bp fragment followed by Sanger sequencing.

#### Biosafety Evaluation of ^68^Ga-NOTA-1A12

To evaluate the biosafety of the ⁶⁸Ga-NOTA-1A12 probe, C57BL/6 mice were randomly divided into two groups: the experimental group received a single intravenous injection of ^68^Ga-NOTA-1A12 at a dose more than 100-fold higher than the equivalent clinical dose (approximately 37 MBq per mouse), while the control group was injected with an equal volume of sterile saline. Body weights were recorded daily from day 0 to day 7 post-injection. On day 7, mice were euthanized, and major organs including the heart, liver, spleen, lung, and kidney were harvested, fixed in 4% paraformaldehyde (Biosharp, China), embedded in paraffin, sectioned, and subjected to hematoxylin and eosin (H&E) staining (Servicebio, China). Histopathological examinations were performed under light microscopy (Leica, Germany) to assess potential structural or morphological differences between groups.

#### AAV-Mediated DDR2 Overexpression in Mouse Lung

To establish a lung-specific DDR2 overexpression model, an adeno-associated virus encoding murine DDR2 (AAV9 [ssAAV.CAG.mDdr2-3FLAG.WPRE.SV40pA]; Guangzhou Paizhen Biotechnology Co., Ltd., Guangzhou, China) was used. For administration, mice were anesthetized with 3% isoflurane and maintained under 1.5% isoflurane during the procedure. After endotracheal intubation, 50 μL of the AAV-DDR2 viral suspension was instilled intratracheally using a microsyringe to ensure uniform delivery into both lungs. Control mice received an equal volume of sterile saline. After instillation, animals were maintained under standard SPF housing conditions for 3-4 weeks to allow sufficient viral expression. Successful DDR2 overexpression was confirmed by PET/CT imaging using the ^68^Ga-NOTA-1A12 tracer and by immunofluorescence staining of lung tissue sections.

#### In Vivo Fluorescence Imaging

Mice were intravenously injected with 15 μg of the fluorescently labeled hFAP antibody and anesthetized with isoflurane. After hair removal, fluorescence images of the lesion sites were acquired using the AniView600 Multi-Mode In Vivo Animal Imaging System equipped with an X-ray imaging module (Guangzhou Biolight Biotechnology Co., Ltd., China). Untreated C57BL/6J mice were used as controls. Regions of interest (ROI) were manually outlined, and the average fluorescence radiance within each ROI was quantified using AniView 100 software (Guangzhou Biolight Biotechnology Co., Ltd., China). The same imaging protocol was also applied to detect fluorescence signal positions on SDS-PAGE gels and to evaluate the uptake of the hFAP-Cy7 probe in 293T and 293T-hFAP+ cells.

#### PET/CT Imaging

PET/CT imaging of mice was performed using a small-animal PET/CT scanner (MadicLab PSA094; Shandong Madic Technology Co., Ltd., China). Images were acquired 40-60 min after intravenous injection of approximately 7.4 MBq of either ^18^F-FAPI or ^68^Ga-NOTA-1A12, while mice were maintained under 1.5% isoflurane anesthesia. For non-human primate imaging, PET/CT scans were conducted using a total-body PET/CT scanner (uEXPLORER; United Imaging Healthcare, Shanghai, China). Blocking experiments were performed by administering an intravenous injection of unlabeled 1A12 nanobody (500 μg per mouse) 8 h prior to the injection of ^68^Ga-NOTA-1A12.The macaque received an intravenous injection of approximately 74 MBq of ^18^F-FAPI on day 4 and an equivalent dose of ^68^Ga-1A12 on day 6 after bleomycin (BLM) administration. Twelve hours post-injection, the animal was anesthetized, and PET/CT imaging was performed.

The clinical PET/CT imaging study using the ^68^Ga-1A12 tracer was approved by the Ethics Committee of Guangdong Provincial Hospital of Chinese Medicine (approval ZF2025-093-01), and written informed consent was obtained from the participating patient. A single patient clinically diagnosed with interstitial lung disease (ILD) underwent PET/CT imaging to evaluate in vivo DDR2-targeted molecular visualization. The patient received an intravenous bolus injection of approximately 2.1-3.5 MBq/kg of ⁶⁸Ga-1A12, followed by whole-body PET/CT acquisition at one hour post-injection using a total-body PET/CT scanner (uEXPLORER, United Imaging Healthcare, Shanghai, China). A low-dose CT scan was obtained for anatomical localization and attenuation correction, after which PET data were collected in list-mode and reconstructed using the ordered-subset expectation maximization (OSEM) algorithm. PET/CT image processing, fusion, and quantitative analysis were performed on a Siemens post-processing workstation, and the standardized uptake value (SUV) of lung lesions was calculated. The resultant PET/CT images, including axial, coronal, sagittal, and maximum-intensity-projection (MIP) views, were further analyzed using PMOD software (version 4.206, PMOD Technologies Ltd., Zurich, Switzerland) to assess the in vivo distribution and uptake characteristics of the ^68^Ga-NOTA-1A12 tracer.

#### PET/CT Image Analysis

PET images were reconstructed using a three-dimensional ordered subset expectation maximization (3D OSEM) algorithm, with CT data applied for attenuation correction and anatomical reference. All imaging data were processed and analyzed using PMOD software (version 4.206; PMOD Technologies Ltd., Zurich, Switzerland). Images were displayed in coronal, sagittal, and transverse planes. The average radioactivity concentration (kBq/cc) within each region was normalized to the injected dose and body weight of the mouse, and expressed as the standardized uptake value (SUV). The lung region was delineated based on CT density values, and Maximum Intensity Projection (MIP) was used to overlay the PET signal for 3D reconstruction of the whole-lung PET image. Calculation of total standardized uptake value (SUV total) was performed as previously described (PMID: 35984444). Briefly, the mean standardized uptake value (SUV mean) of the mediastinal blood pool was used as the absolute threshold. The region of interest (ROI) was determined by three-dimensional segmentation of all voxels above this threshold, and the total volume of the ROI was defined as the total active volume. The SUV total of the ^68^Ga-NOTA-1A12 tracer in the lung was calculated by multiplying the SUV mean of the ROI by the total active volume.

#### Micro-CT and 3D Reconstruction of Pulmonary Ventilation Regions

Micro-CT imaging was performed and analyzed as previously described (PMID: 34561295). Briefly, mice were anesthetized with isoflurane and scanned using a high-resolution micro-CT system (Super Nova CT, SNC-100; PINGSENG Healthcare, Kunshan, Jiangsu, China) with a voxel size of 50 µm. The pulmonary ventilation regions were reconstructed and analyzed according to the manufacturer’s instructions using the AVATAR 1.5.0 three-dimensional (3D) finite element analysis software (PINGSENG Healthcare).

#### Western blotting

Protein expression was analyzed by western blotting. Briefly, the tissues or cells were lysed using ice-cold radioimmunoprecipitation assay lysis buffer (RIPA, ZHHC Biotechnology Co., Ltd, Shaanxi, China) with addition of 1% phenylmethanesulfonyl fluoride (PMSF, ZHHC Biotechnology Co., Ltd, Shaanxi, China) in a high-throughput tissue grinder (KeCheng, KC-48, Hunan, China). After centrifuged, the supernatant was collected and the total protein concentration was detected by bicinchoninic acid protein assay kit (BCA, Biosharp, Anhui, China). Protein was separated using SDS-PAGE and then transferred onto a poly (vinylidene fluoride) membrane (PVDF, Merck Millipore, Germany). After blocked in 5 % skim milk (Bioforxx, Germany) at room temperature for 1 hour, the membrane was incubated with primary antibody at 4℃ overnight and then incubated with horseradish peroxidase (HRP)-conjugated secondary antibody at room temperature for 1 h. Finally, the specific protein band was visualized using an ultra-sensitive ECL chemiluminescent substrate (Biosharp, Anhui, China) in the GelView 6000 Pro Multifunctional Imaging Station (Biolight Biotechnology Co., Ltd, Guangdong, China). The primary antibodies used in this study were as follows: anti-DDR2 (R&D Systems, AF2538, 1:2000); anti-hFAP (Cell Signaling Technology, #66562,USA) ; anti-β-actin (Yeasen, 30102ES60, 1:1000).

#### Immunofluorescence Staining

Lung tissues were fixed in 4% paraformaldehyde, embedded in paraffin, and sectioned for mounting. The sections were deparaffinized with two changes of xylene (20 min each), rehydrated through a graded ethanol series, and subjected to antigen retrieval by boiling in retrieval buffer for 15 min, followed by cooling to room temperature. After three washes with PBS, the sections were blocked with 5% bovine serum albumin (BSA, ) for 1 h and incubated with primary antibodies overnight at 4 °C. After washing, the sections were incubated with species-appropriate Alexa Fluor-conjugated secondary antibodies for 2 h at room temperature, followed by nuclear staining with 4′,6-diamidino-2-phenylindole (DAPI; Thermo Fisher, 1 µg/mL). The following primary antibodies were used: anti-DDR2 (R&D Systems, AF2538; 1:50) and anti-FAP (Abmart, M079003; 1:500). Secondary antibodies included Alexa Fluor 488/594 labeled anti-mouse/goat IgG (Abcam; 1:200). Whole-slide fluorescence images were captured using an Aperio CS2 Whole Slide Scanner (Leica, Nussloch, Germany) or a fluorescence microscope. The expression areas of DDR2 and FAP, as well as their co-localization patterns, were analyzed using ImageJ software (Fiji version, https://imagej.net/Fiji).

#### Flow Cytometry

Flow cytometry was performed to analyze DDR2 and FAP expression in lung tissue from patients with idiopathic pulmonary fibrosis (IPF). Fresh IPF lung tissues were enzymatically dissociated into single-cell suspensions according to standard procedures. The cell suspensions were filtered through a 70 μm cell strainer (BD Biosciences, 352350, USA) and washed twice with phosphate-buffered saline (PBS) containing 1% bovine serum albumin (BSA). The cells were then incubated with fluorescence-conjugated antibodies, including anti-DDR2/AF488 and anti-FAP/APC, for 30 min at 4 °C in the dark. After staining, cells were washed twice with PBS and resuspended for acquisition. Flow cytometric analysis was performed using a BD LSRFortessa™ flow cytometer (BD Biosciences, USA), and data were analyzed using FlowJo software (version 10.8, BD Biosciences). The proportion of DDR2⁺, FAP⁺, and DDR2⁺FAP⁺ cell populations was determined based on fluorescence intensity compared with unstained and single-stained controls. Similarly, to confirm hFAP overexpression in 293T-hFAP+ cells, wild-type (WT) and hFAP+ 293T cells were first incubated with a primary anti-hFAP antibody, followed by an anti-His APC secondary antibody recognizing the His tag on the anti-hFAP antibody. The fluorescence signal was measured by flow cytometry.

### Statistical analysis

The data analysis was performed using GraphPad Prism V.8 (GraphPad Software, La Jolla, California). All results were presented as mean ± standard deviation. The significant difference between experimental groups was analyzed using one-way analysis of variance (ANOVA). The *p* value < 0.05 was considered as statistically significant.

