## Supplemental Figures for "A DDR2-targeted PET tracer images activated fibroblasts in early pulmonary fibrosis"

**Figure. S1 Correlation of DDR2 with fibrosis-related genes and co-localization analysis of DDR2 and FAP in IPF lung tissues. a**, Correlation analysis of *DDR2* with *TGFB1, COL1A1,* and *COL1A2* expression in fibroblast and myofibroblast clusters (GSE136831). *DDR2* expression was positively correlated with *TGFB1* ($R = 0.33$, $P < 0.0001$) and negatively correlated with *COL1A1* and *COL1A2* ($R = -0.24$ for both, $P < 0.0001$). **b**, Representative high-magnification immunofluorescence images showing co-localization (yellow) of DDR2 (red) and FAP (green) signals in fibrotic regions of IPF lung tissues, primarily along the cell membrane. **c**, Quantitative analysis of co-localization areas showing that DDR2-positive regions occupied a larger proportion of the fibrotic area compared with FAP, with an overlapping co-localized fraction of 6.37%.

**Figure. S2 Purity characterization of 1A12 nanobody and validation of DDR2 overexpression in 293T cells.**

**a**, Ultra-performance liquid chromatography (UPLC) and time-of-flight mass spectrometry (TOF-MS) analyses confirming the high purity and molecular weight of the 1A12 nanobody. UPLC showed a single main peak with a retention time of 3.62 min, and TOF-MS identified a dominant peak corresponding to a molecular weight of 15,890 Da, consistent with the theoretical value of 1A12. **b,** Biolayer Interferometry analysis to determine the affinity of 1A12 binding to DDR2. **c**, Flow cytometry analysis demonstrating successful establishment of DDR2-overexpressing 293T (DDR2+) cells, with 82.08% DDR2-positive population compared to 0.90% in wild-type (WT) cells. **d**, Western blot analysis further confirming high DDR2 protein expression in DDR2+ cells, while WT cells showed minimal background signal. β-actin served as a loading control.

**Figure. S3 In vivo safety assessment of the $^{68}$Ga-1A12 probe in mice. a**, Body weight monitoring of C57BL/6 mice for seven consecutive days after intravenous administration of a high dose of $^{68}$Ga-NOTA-1A12 (approximately 37 MBq per mouse, >100-fold higher than the clinical imaging dose). Both the $^{68}$Ga-NOTA-1A12 treated group and the control group (saline) showed a gradual increase in body weight with no significant difference between groups (ns: not significant, $n = 5$, one-way ANOVA). **b**, Representative hematoxylin-eosin (H&E) staining of major organs, including the heart, lung, liver, kidney, and spleen, showing normal histological architecture with no evidence of tissue injury or inflammation in the $^{68}$Ga-NOTA-1A12 treated group compared with the control group.

**Figure. S4 Validation of the hFAP knock-in (hFAP-KI) mouse model and in vitro/in vivo characterization of the hFAP-Cy7 probe.** **a**, Western blot analysis confirming successful human FAP (hFAP) knock-in expression in lung tissues of hFAP-KI mice. hFAP protein levels were markedly elevated after bleomycin (BLM) treatment, whereas WT mice showed no detectable hFAP expression. β-actin served as a loading control. **b**, SDS-PAGE analysis of purified Cy7 dye, hFAP antibody, and the Cy7-conjugated hFAP antibody (hFAP-Cy7), demonstrating a clear molecular band at about 30 kDa for hFAP-Cy7, consistent with successful dye conjugation. **c**, Fluorescence gel imaging confirmed strong Cy7 fluorescence associated with the hFAP-Cy7 band, while free Cy7 dye showed a lower-molecular-weight signal, further validating proper probe assembly. **d**, Flow cytometry analysis of hFAP-overexpressing 293T cells (hFAP+) labeled with anti-His-APC antibody showed a distinct positive population (54.51%), whereas WT 293T cells displayed minimal background signal (0.99%), confirming specific recognition of hFAP. **e**, *In vitro* fluorescence imaging of WT and hFAP+ 293T cells incubated with hFAP-Cy7 probe. Strong Cy7 fluorescence was observed only in the hFAP+ group, demonstrating selective probe binding. **f**, Micro-CT airway reconstruction and in vivo fluorescence imaging of BLM-treated WT and hFAP-KI mice. Airway 3D modeling revealed pronounced fibrotic remodeling in the left lung of BLM-treated hFAP-KI mice. Correspondingly, hFAP-Cy7 imaging showed clear fluorescence accumulation in the fibrotic region of hFAP-KI mice, with no detectable signal in WT mice. **g**, PET/CT imaging of the same animals using the $^{18}$F-FAPI probe. BLM-treated hFAP-KI mice exhibited strong radiotracer uptake localized to the fibrotic lung, whereas WT mice showed negligible uptake. Axial and coronal CT and PET/CT fusion images corroborate successful model establishment and demonstrate that $^{18}$F-FAPI selectively targets hFAP-expressing fibrotic lesions.

**Figure. S5 Three-Dimensional (3D) PET imaging Reconstruction and SUV total Analysis of $^{68}$Ga-NOTA-1A12 in the Unilateral Pulmonary Fibrosis Model.** **a**, Representative PET/CT 3D reconstructions and fused PET/CT images of C57BL/6 mice at baseline (Con) and at days 3, 7, 14, 21, and 28 following unilateral intratracheal bleomycin (BLM) administration. A progressive increase in radiotracer accumulation was observed in the left lung during early fibrotic stages (days 3~14), with peak uptake at day 14, followed by a gradual decline during later stages (days 21 and 28). In both 3D reconstruction and PET/CT fusion images, the dotted outlines denote the segmented lung region. **b**, Quantitative SUV total analysis of $^{68}$Ga-NOTA-1A12 uptake in the left lung across time points ($n = 8$ per group). SUV total increased significantly from day 7 onward, peaked on day 14 ($2.85 \pm 0.48$), and decreased during later fibrotic stages, consistent with disease progression. Statistical comparisons were performed using one-way ANOVA (****$P < 0.0001$, ns: not significant).

**Figure. S6 Validation of anti-mouse FAP antibody specificity in MC38 cells.** **a**, Representative immunofluorescence staining of murine colon carcinoma MC38 cells (MC38) and MC38 cells stably overexpressing murine FAP (MC38-mFAP). In MC38-mFAP cells, strong membrane-associated green fluorescence signals corresponding to FAP staining were observed, while wild-type MC38 cells displayed only weak background fluorescence. Nuclei were counterstained with DAPI (blue).

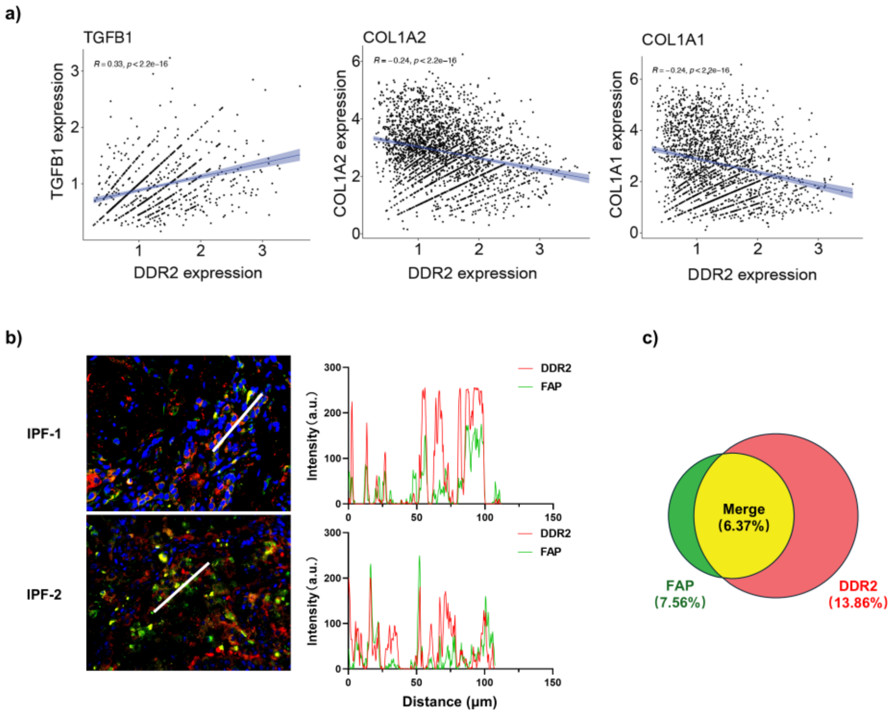
a)
TGFB1
COL1A2
COL1A1
TGFB1 expression
COL1A2 expression
COL1A1 expression
DDR2 expression
DDR2 expression
DDR2 expression
b)
IPF-1
IPF-2
DDR2
FAP
Intensity (a.u.)
Distance (μm)
c)
Merge
(6.37%)
FAP
(7.56%)
DDR2
(13.86%)

### Figure. S1

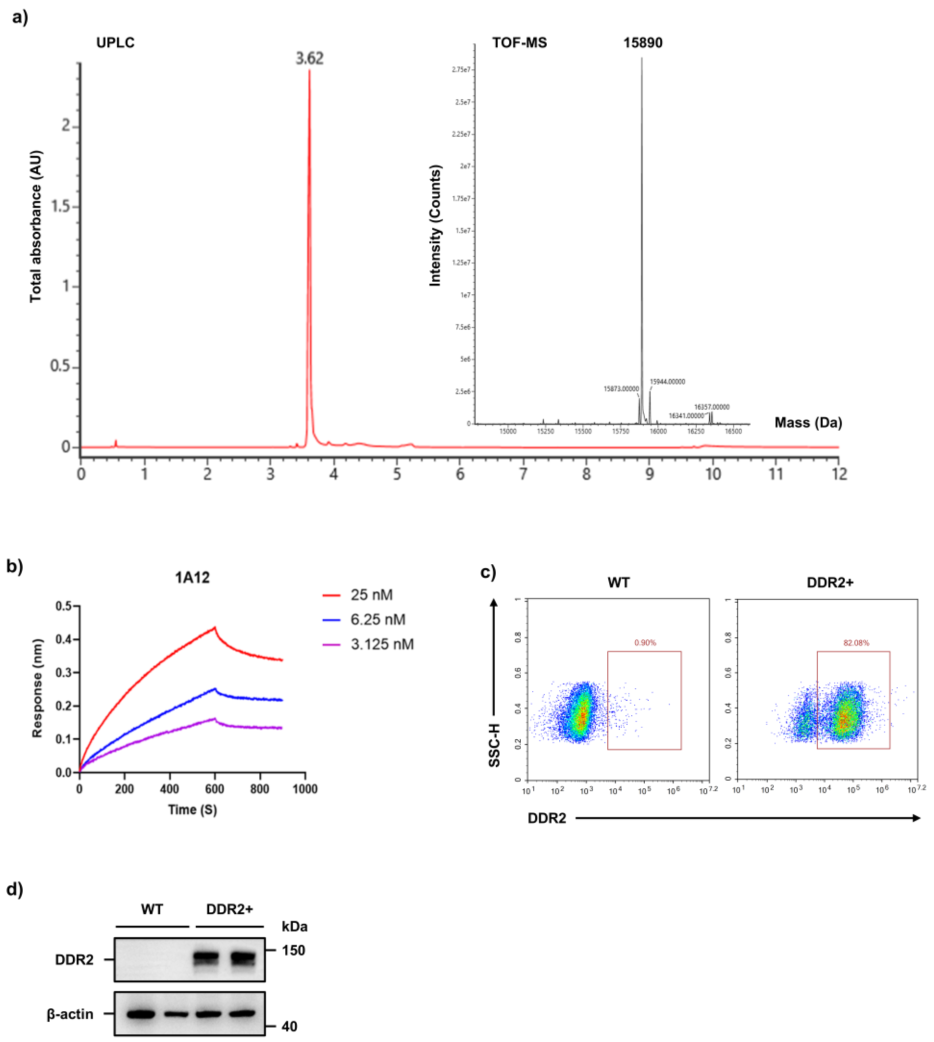
a)
UPLC
3.62
Total absorbance (AU)
TOF-MS
15890
Intensity (Counts)
15873.00000
15944.00000
16341.00000
16357.00000
Mass (Da)
b)
1A12
25 nM
6.25 nM
3.125 nM
Response (nm)
Time (S)
c)
WT
DDR2+
0.90%
82.08%
SSC-H
DDR2
d)
WT
DDR2+
kDa
DDR2
150
β-actin
40

Figure. S2

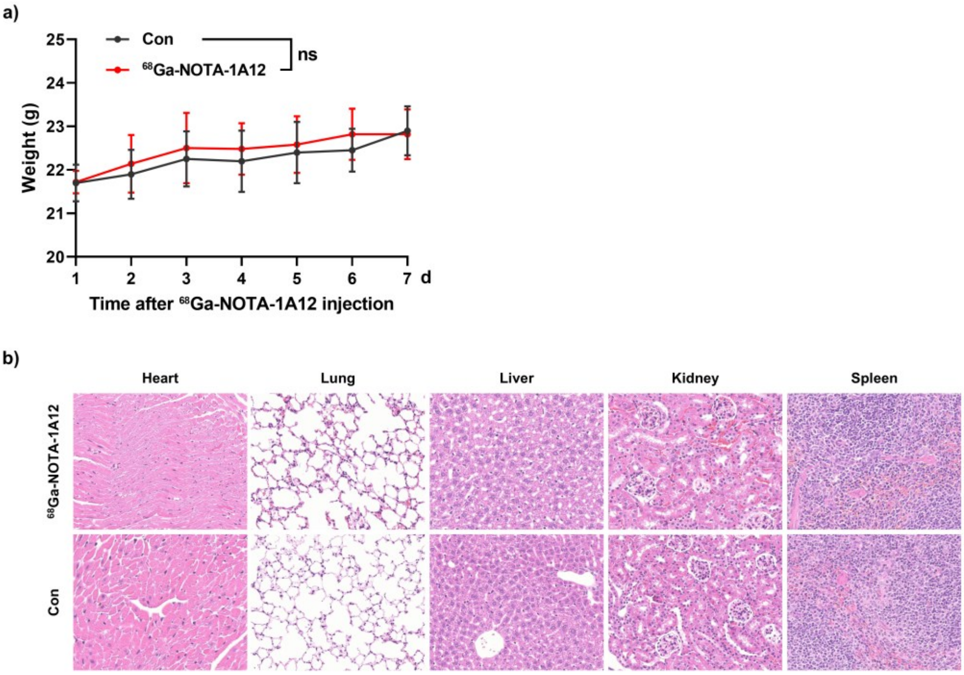
a)
Con
68Ga-NOTA-1A12
ns
Weight (g)
25
24
23
22
21
20
1
2
3
4
5
6
7
d
Time after 68Ga-NOTA-1A12 injection
b)
Heart
Lung
Liver
Kidney
Spleen
68Ga-NOTA-1A12
Con

**Figure. S3**

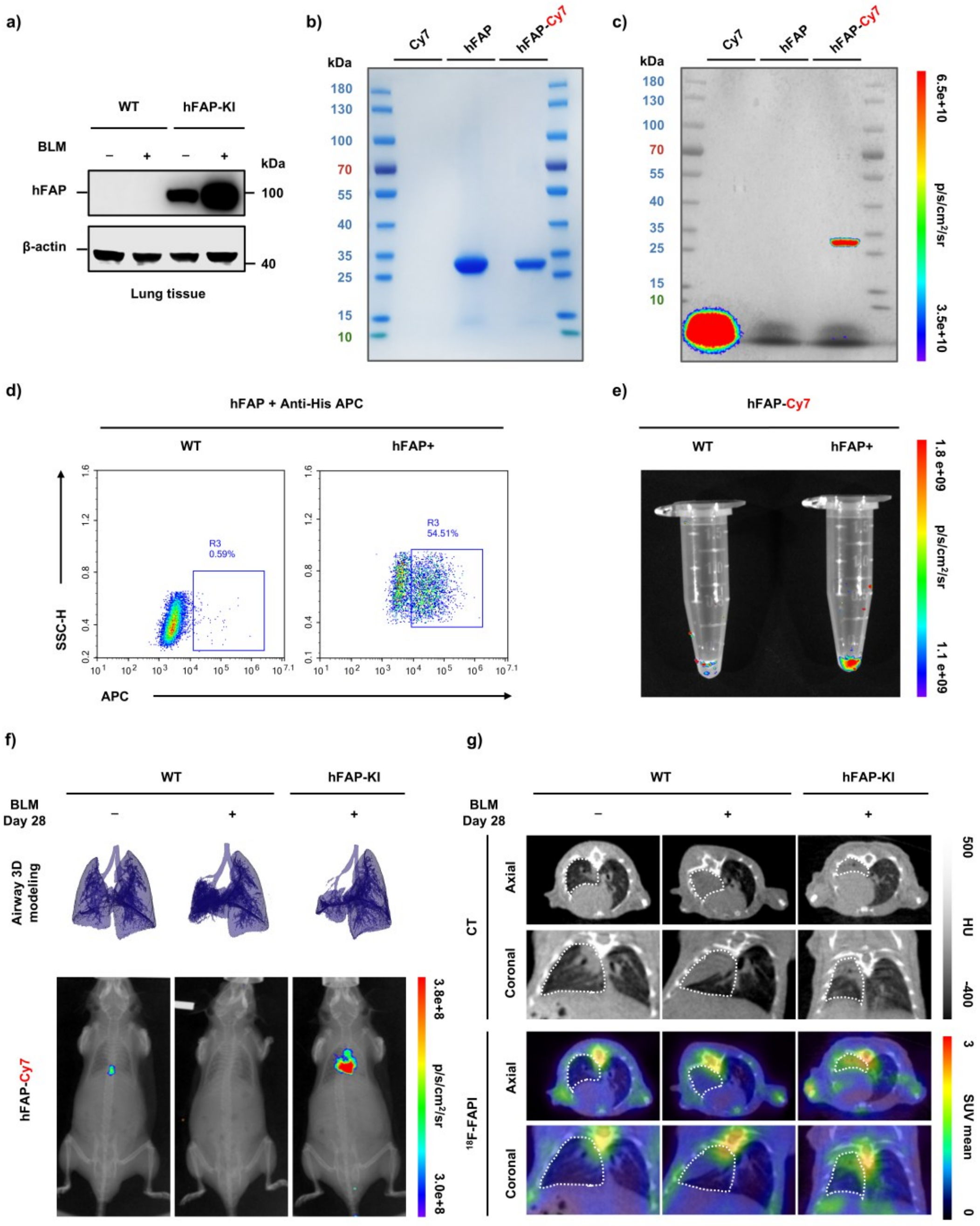
a)
WT
hFAP-KI
BLM
kDa
hFAP
100
β-actin
40
Lung tissue
b)
Cy7
hFAP
hFAP-Cy7
kDa
180
130
100
70
55
40
35
25
15
10
c)
Cy7
hFAP
hFAP-Cy7
kDa
6.5e+10
p/s/cm²/sr
3.5e+10
d)
hFAP + Anti-His APC
WT
hFAP+
R3
0.59%
R3
54.51%
SSC-H
APC
e)
hFAP-Cy7
WT
hFAP+
1.8 e+09
p/s/cm²/sr
1.1 e+09
f)
WT
hFAP-KI
BLM
Day 28
Airway 3D modeling
hFAP-Cy7
3.8e+8
p/s/cm²/sr
3.0e+8
g)
WT
hFAP-KI
BLM
Day 28
CT
Axial
Coronal
500
HU
-400
18F-FAPI
3
SUV mean
0

**Figure. S4**

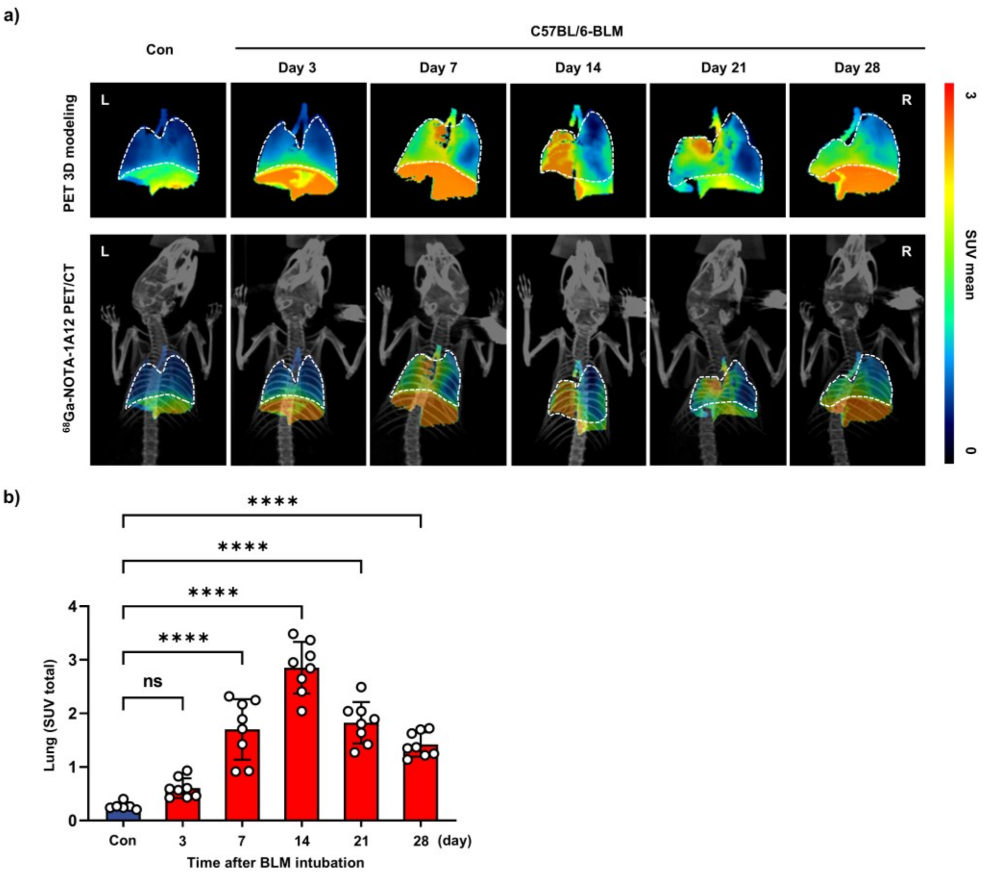
a)
Con
C57BL/6-BLM
Day 3
Day 7
Day 14
Day 21
Day 28
PET 3D modeling
68Ga-NOTA-1A12 PET/CT
L
R
SUV mean
3
0
b)
****
****
****
****
ns
Lung (SUV total)
4
3
2
1
0
Con
3
7
14
21
28
(day)
Time after BLM intubation

**Figure. S5**

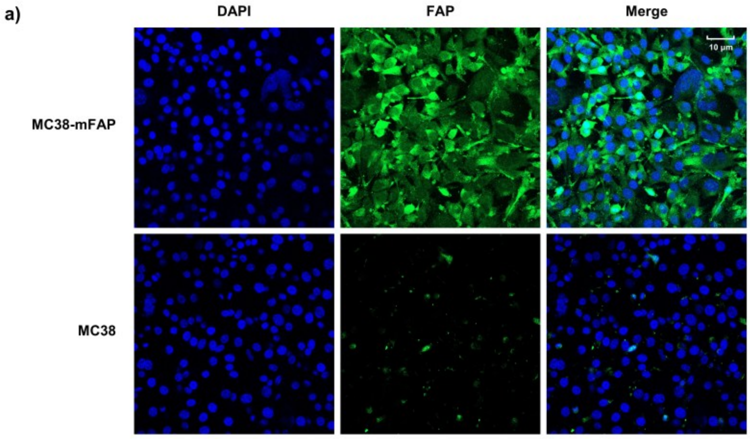


Figure. S6
